# Dominance of Alpha and Iota variants in SARS-CoV-2 vaccine breakthrough infections in New York City

**DOI:** 10.1101/2021.07.05.21259547

**Authors:** Ralf Duerr, Dacia Dimartino, Christian Marier, Paul Zappile, Guiqing Wang, Jennifer Lighter, Brian Elbel, Andrea Troxel, Adriana Heguy

## Abstract

The efficacy of COVID-19 mRNA vaccines is high, but breakthrough infections still occur. We compared the SARS-CoV-2 genomes of 76 breakthrough cases after full vaccination with BNT162b2 (Pfizer/BioNTech), mRNA-1273 (Moderna), or JNJ-78436735 (Janssen) to unvaccinated controls (February-April 2021) in metropolitan New York, including their phylogenetic relationship, distribution of variants, and full spike mutation profiles. Their median age was 48 years; seven required hospitalization and one died. Most breakthrough infections (57/76) occurred with B.1.1.7 (Alpha) or B.1.526 (Iota). Among the 7 hospitalized cases, 4 were infected with B.1.1.7, including 1 death. Both unmatched and matched statistical analyses considering age, sex, vaccine type, and study month as covariates supported the null hypothesis of equal variant distributions between vaccinated and unvaccinated in chi-squared and McNemar tests (p>0.1) highlighting a high vaccine efficacy against B.1.1.7 and B.1.526. There was no clear association among breakthroughs between type of vaccine received and variant. In the vaccinated group, spike mutations in the N-terminal domain and receptor-binding domain that have been associated with immune evasion were overrepresented. The evolving dynamic of SARS-CoV-2 variants requires broad genomic analyses of breakthrough infections to provide real-life information on immune escape mediated by circulating variants and their spike mutations.

## INTRODUCTION

The novel betacoronavirus SARS-CoV-2 arose as a new human pathogen at the end of 2019, and rapidly spread to every corner of the globe, causing a pandemic of enormous proportions, with 179 million cumulative infections and almost 4 million deaths counted worldwide as of mid-June 2021 (10). As soon as the causative agent was identified and sequenced (11), massive efforts to develop vaccines were initiated and tested simultaneously in multiple clinical trials.

These efforts led to the rapid deployment of several of these vaccines by December 2020, less than a year after the first SARS-CoV-2 viral sequence had been determined. In the United States, vaccination started in December 2020, using two novel mRNA-based vaccines, BNT162b2 (Pfizer/BioNTech), and mRNA-1273 (Moderna), both using the SARS-CoV-2 spike mRNA sequence of the original Wuhan variant. The Janssen COVID-19 vaccine, JNJ-78436735, was also deployed shortly after the mRNA vaccines, and these vaccines were shown to have high efficacy in clinical trials (12-14). With millions of people vaccinated in multiple countries, they have proven to be highly effective in the real world (15-21), although a very small percentage of breakthrough infections have occurred, as expected (8, 22-31).

Contemporaneously with the vaccination efforts in several countries, new SARS-CoV-2 variants emerged, four of which were designated by the WHO as variants of concern (VOC), B.1.1.7 or Alpha, B.1.351 or Beta, P.1 or Gamma, and B.1.617.2 or Delta, which arose originally in the UK, South Africa, Brazil, and India, respectively (3, 6). VOCs are classified as such if they are more transmissible, cause more severe disease or a significant reduction in neutralization by antibodies generated via previous infection or after vaccination. In addition, there are variants of interest (VOI), which carry mutations that have been associated with changes to receptor binding, reduced neutralization by anti-SARS-CoV-2 antibodies, or may increase transmissibility or severity. One of these variants is B.1.526 (Iota), which arose in New York City in late December 2020 (32, 33). Although it is not yet clear that any particular VOC or VOI is associated with vaccine breakthrough, data from Israel and Washington State suggest that there might be higher vaccine breakthrough rates with VOCs (8, 9).

In addition, in vitro selection, deep-mutational scanning of spike libraries, yeast display, epidemiological and structural studies have revealed critical spike mutations that can escape monoclonal antibodies and convalescent sera (5, 7, 34-38). Thus, it is possible that certain amino acid mutations in spike, irrespective of affiliation to a specific VOC or VOI, are critical for vaccine breakthgh, but this has only been rudimentarily studied (9, 26). There is scarcity of data about determinants of vaccine breakthrough. The few reported studies have included breakthrough cases after first or second immunization, however, breakthrough cases after full vaccination remained moderate or low (8, 22-31). Thus, comprehensive studies with site-specific mutation analyses are needed on a larger number of fully vaccinated individuals from different geographic regions. Here, we added such data from the NY metropolitan area. We carried out full SARS-CoV-2 genome sequencing of SARS-CoV-2-positive individuals, 14 days after their completed vaccination series with mostly Pfizer and Moderna vaccines in the multi-center NYU Langone Health System. Analyses included statistical comparison of variant distribution as well as mutation rates at every residue in spike between vaccinated and unvaccinated individuals.

## RESULTS

### Alpha and Iota variants dominate vaccine breakthrough infections in metropolitan New York

A total of 126,367 fully vaccinated individuals were recorded in our electronic health records by April 30^th^, 2021, of whom the majority (123,511, 98%) were vaccinated with mRNA vaccines (Pfizer/BioNTech = 103,166 and Moderna = 20,345), and the rest (2,856) with the adenovirus-based Janssen COVID-19 vaccine, administered as single dose. We recorded 101 cases of vaccine breakthrough infection (77x Pfizer/BioNTech, 17x Moderna, and 7x Janssen) between February 1^st^ and April 30^th^ of 2021, representing 1.4% of the 7147 total SARS-CoV-2 positive cases in our healthcare system and 0.08% of the fully vaccinated population in our medical records. Out of the 101 cases, 76 cases (75%) yielded full SARS-CoV2 genomes (61x Pfizer/BioNTech, 11x Moderna, and 4x Janssen) that passed quality control (QC) and allowed us to determine the Pango lineage and mutations across the viral genome, including the spike gene. The median Ct value for the 101 breakthroughs was 27 (range 13-42). As expected, the 25 excluded breakthrough cases with low genome coverage and failed QC had significantly higher cycle threshold (Ct) values (median: 34, range: 27-42) than the 76 samples with full viral genome coverage (P<0.0001, Mann Whitney test), which had Ct values below 36 (median: 24, range: 13-36) (**Figure 1A**).

**Figure 1.**
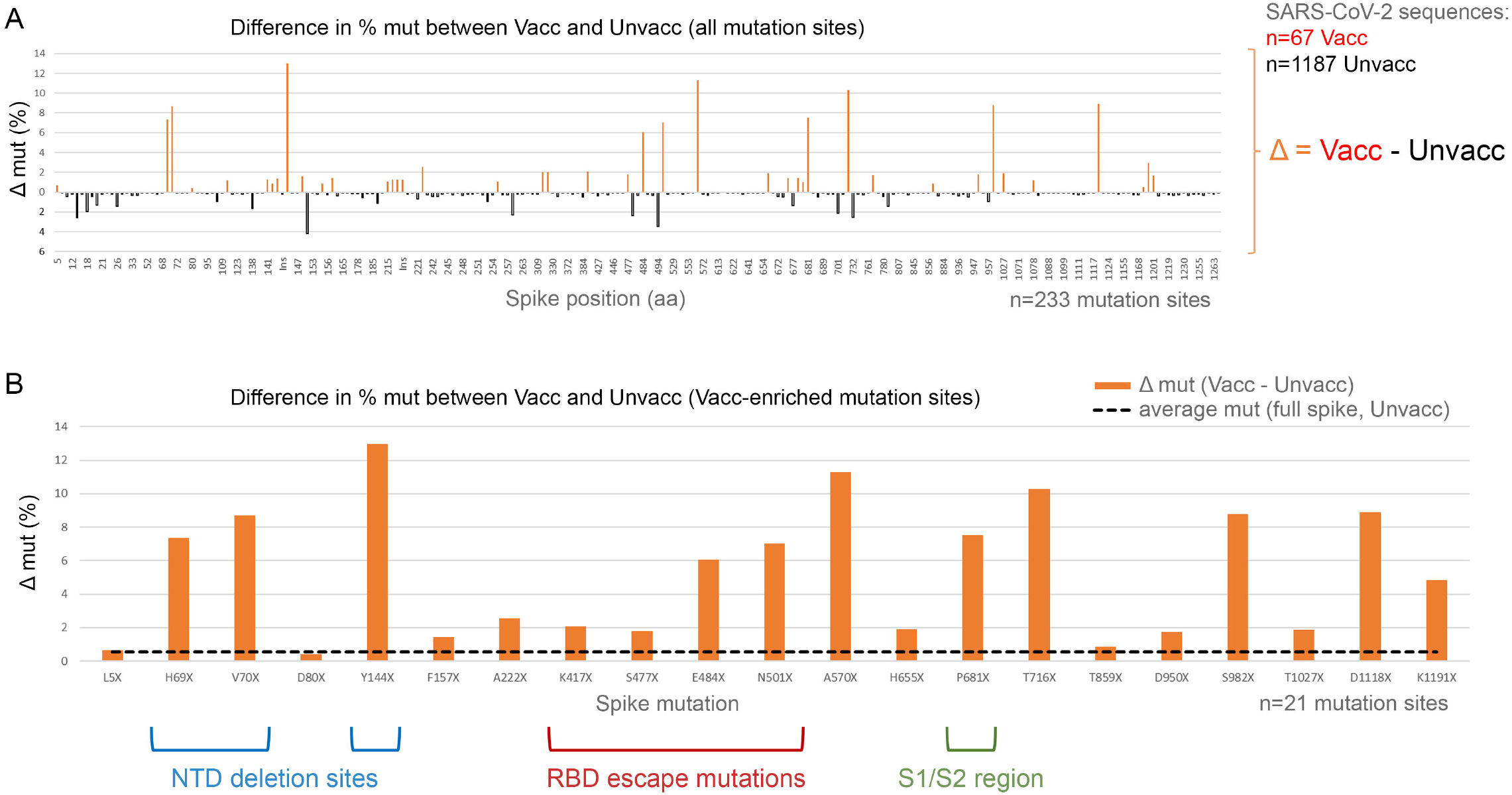
RT-qPCR Ct values in post-vaccine breakthrough infections. **A)** Ct plots of samples that yielded a full genome with sufficient coverage to determine lineage and mutations (QC passed = >23,000 bp and >4000X coverage; QC failed = <23,000 bp and <4000X coverage). **B)** Ct plots of all samples that passed QC, by lineages, B.1.1.7 (Alpha) and B.1.526 (Iota), compared to all others. Hospitalized cases are shown in larger sized black symbols. QC: quality control.

Although B.1.1.7 (Alpha) infection has been associated with overall higher viral load/lower Ct values (39, 40), the Ct values in our recorded breakthrough infections were not significantly different for B.1.1.7 compared to B.1.526 (Iota) or all other variants (**Figure 1B**). Among the 76 COVID-19 cases post-vaccination with adequate SARS-CoV-2 genome coverage, the median age was 48 years, 37 were male and 39 were female. Seven required hospitalization for COVID-19, among whom there was one death in an elderly patient with multiple comorbidities who already was on in-home oxygen previous to post-vaccination COVID-19 infection and had a lengthy stay at the ICU (**Tables 1 and 2, Supplementary Table 3**).

**Table 1.** Vaccine breakthrough study population (<60 days after completion of vaccination)

| Sample ID | Sex | Age Range | Vaccine | Days post completion of vaccination | Hospitalization | Ct | Pango Lineage |
| --- | --- | --- | --- | --- | --- | --- | --- |
| P21-2331 | F | 81-90 | Pfizer | 17 | Y (COVID) | 23 | B.1.526 |
| P21-2383 | M | 41-50 | Pfizer | 17 | N | 33 | B.1.526 |
| P21-1948 | F | 81-90 | Pfizer | 18 | Y (not COVID) | 14 | B.1.1.519 |
| P21-1402 | F | 51-60 | Janssen | 20 | Y (COVID) | 25 | B.1 |
| P21-2378 | M | 61-70 | Pfizer | 20 | N | 18 | B.1.1 |
| P21-1234 | M | 71-80 | Pfizer | 21 | N | 25 | B.1.526 |
| P21-1038 | F | 71-80 | Moderna | 22 | N | 27 | B.1.2 |
| P21-1574 | F | 21-30 | Pfizer | 22 | N | 23 | B.1 |
| P21-2192 | M | 41-50 | Janssen | 23 | Y (COVID) | 25 | B.1.1.7 |
| P21-2580 | M | 61-70 | Pfizer | 23 | N | 27 | B.1.2 |
| P22-1140 | M | 61-70 | Pfizer | 23 | Y (COVID) | 15 | B.1.526.2 |
| P21-1989 | M | 21-30 | Janssen | 26 | N | 25 | B.1.526 |
| P21-2382 | M | 61-70 | Pfizer | 26 | N | 20 | B.1.1.7 |
| P21-2376 | F | 21-30 | Pfizer | 27 | N | 19 | B.1.526.1 |
| P21-2764 | F | 51-60 | Pfizer | 27 | Y (COVID) | 31 | B.1.1.7 |
| P21-0524 | M | 21-30 | Pfizer | 28 | N | 33 | B.1.526.2 |
| P21-2765 | M | 81-90 | Moderna | 28 | N | 17 | B.1.526 |
| P21-2388 | M | 31-40 | Pfizer | 31 | N | 18 | A.2.5.2 |
| P21-1292 | F | 71-80 | Pfizer | 33 | Y (COVID) | 20 | B.1.1.7 |
| P21-1296 | F | 41-50 | Pfizer | 40 | N | 31 | B.1.526 |
| P21-1415 | M | 61-70 | Moderna | 42 | N | 24 | B.1.1.7 |
| P21-2563 | F | 61-70 | Janssen | 42 | Y (not COVID) | 23 | B.1.429 |
| P21-1416 | F | 41-50 | Pfizer | 45 | N | 27 | B.1.2 |
| P21-2370 | M | 71-80 | Pfizer | 48 | N | 31 | B.1.526 |
| P21-1407 | M | 31-40 | Pfizer | 49 | N | 28 | B.1.1.519 |
| P21-2738 | M | 71-80 | Moderna | 49 | N | 24 | B.1.1.7 |
| P21-1993 | M | 81-90 | Moderna | 50 | Y (COVID) | 23 | B.1.1.7 |
| P21-2384 | F | 71-80 | Moderna | 50 | N | 24 | B.1.1.7 |
| P21-2368 | F | 21-30 | Pfizer | 52 | N | 22 | B.1.1.7 |
| P21-1595 | F | 71-80 | Pfizer | 53 | N | 18 | B.1.1.7 |
| P21-0852 | F | 21-30 | Pfizer | 54 | N | 22 | B.1.1.7 |
| P21-1411 | F | 51-60 | Pfizer | 54 | N | 30 | B.1.111 |
| P21-2371 | F | 61-70 | Moderna | 54 | N | 20 | B.1.429 |
| P21-2143 | F | 81-90 | Pfizer | 55 | Y (not COVID) | 20 | B.1.526 |
| P21-1596 | M | 71-80 | Moderna | 57 | N | 34 | B.1.1 |
| P21-2365 | F | 21-30 | Pfizer | 57 | N | 27 | B.1.526.1 |
| P21-2366 | M | 71-80 | Pfizer | 57 | N | 24 | B.1.526.2 |
| P21-1293 | F | 31-40 | Pfizer | 58 | N | 27 | B.1.526.2 |
| P21-1405 | F | 31-40 | Pfizer | 58 | N | 25 | B.1.526 |
| P21-2364 | M | 51-60 | Pfizer | 59 | N | 31 | B.1.1.7 |

Ct=cycle threshold from real-time RT-PCR SARS-CoV-2 test

**Table 2.** Vaccine breakthrough study population (>60 days after completion of vaccination)

| Sample ID | Sex | Age Range | Vaccine | Days post completion of vaccination | Hospitalization | Ct | Pango Lineage |
| --- | --- | --- | --- | --- | --- | --- | --- |
| P21-1362 | F | 41-50 | Pfizer | 61 | N | 20 | B.1.526.1 |
| P21-2057 | M | 71-80 | Pfizer | 61 | N | 16 | B.1.526 |
| P21-2124 | F | 21-30 | Pfizer | 61 | N | 24 | B.1.526 |
| P21-1412 | F | 51-60 | Pfizer | 62 | N | 29 | B.1.526.1 |
| P21-2381 | M | 71-80 | Moderna | 64 | N | 22 | B.1.575 |
| P21-1406 | F | 31-40 | Pfizer | 65 | N | 13 | B.1.526.1 |
| P21-2379 | F | 41-50 | Pfizer | 65 | N | 22 | B.1.1.7 |
| P21-2372 | M | 21-30 | Pfizer | 66 | N | 21 | B.1.526 |
| P21-1413 | M | 31-40 | Pfizer | 67 | N | 17 | B.1.526 |
| P21-2137 | F | 31-40 | Moderna | 67 | N | 18 | B.1.526 |
| P21-2191 | M | 31-40 | Pfizer | 68 | N | 18 | B.1.526 |
| P21-1394 | F | 21-30 | Pfizer | 70 | N | 32 | B.1.1.7 |
| P21-1396 | F | 31-40 | Pfizer | 70 | N | 27 | B.1.1.7 |
| P21-2380 | M | 31-40 | Pfizer | 70 | N | 23 | B.1.526.1 |
| P21-1970 | F | 31-40 | Moderna | 71 | N | 27 | B.1.1.7 |
| P21-1403 | F | 21-30 | Pfizer | 72 | N | 27 | B.1.526 |
| P21-2385 | M | 51-60 | Pfizer | 72 | N | 29 | B.1.1.7 |
| P21-1404 | F | 41-50 | Pfizer | 73 | N | 17 | B.1.1.7 |
| P21-1984 | M | 51-60 | Pfizer | 76 | N | 19 | B.1.526.2 |
| P21-1985 | M | 21-30 | Pfizer | 76 | N | 23 | B.1.1.7 |
| P21-1992 | M | 41-50 | Pfizer | 76 | N | 33 | B.1.1.7 |
| P21-2567 | F | 71-80 | Pfizer | 77 | N | 20 | B.1.1.7 |
| P21-1964 | M | 51-60 | Pfizer | 78 | N | 21 | B.1.1.7 |
| P21-2373 | M | 31-40 | Pfizer | 78 | N | 33 | B.1.1.7 |
| P21-1556 | M | 41-50 | Pfizer | 85 | N | 19 | B.1.526.2 |
| P21-2096 | F | 31-40 | Pfizer | 88 | N | 33 | B.1.575 |
| P21-1976 | M | 31-40 | Pfizer | 89 | N | 22 | B.1.526.1 |
| P21-2577 | F | 31-40 | Pfizer | 92 | N | 25 | B.1.1.7 |
| P21-2193 | F | 41-50 | Pfizer | 94 | N | 25 | P.1 |
| P21-2105 | M | 21-30 | Pfizer | 97 | N | 25 | B.1.1.7 |
| P21-3585 | M | 61-70 | Pfizer | 98 | Y (not COVID) | 36 | AV.1 |
| P21-2592 | F | 21-30 | Pfizer | 99 | N | 25 | B.1.526 |
| P21-2573 | F | 31-40 | Pfizer | 100 | N | 19 | B.1.1.7 |
| P21-2594 | F | 31-40 | Pfizer | 102 | N | 18 | B.1 |
| P21-2375 | M | 51-60 | Pfizer | 105 | N | 30 | B.1.526 |
| P21-2535 | M | 51-60 | Pfizer | 108 | N | 14 | B.1.301 |

The distribution of PANGO lineages in the 76 vaccine breakthroughs was as follows: 26 (34.2%) had B.1.1.7, 31 (40.7%) had B.1.526 (including sublineages B.1.526.1 and B.1.526.2), 1 (1.3%) had P.1, and 18 (23.7%) had other variants (**Supplementary Figure 1**). Among the 1,046 sequences from the group of unvaccinated patients, 304 (29.0%) had B.1.1.7, 423 (40.4%) had B.1.526 (including sublineages B.1.526.1, B.1.526.2, and B.1.526.3), 12 (0.07%) had P.1, and 307 (29.3%) had other variants. Of the seven COVID-19 hospitalizations, four were infected with B.1.1.7, including the fatal case, two with B.1.526, and one with B.1 (containing P681H; not a VOC or VOI). All hospitalizations due to COVID-19 occurred in patients who got infected <60 days (**Table 1**) and none in those >60 days after completion of the vaccination series (**Table 2**).

### VOCs and VOIs are equally distributed in vaccinated and unvaccinated infections

To compare vaccinated breakthrough infections with unvaccinated controls statistically, we included 1,046 unvaccinated individuals from the same study cohort who became SARS-CoV-2-infected in the same study months (February-April 2021) as the vaccine breakthrough cases. A chi-squared test for rejecting the null hypothesis of equal Pango lineage distributions (B.1.1.7, B.1.526, and other variants combined) between vaccinated and unvaccinated patients resulted in a p-value of 0.70.

To address confounding and other sources of bias arising from the use of observational data, we estimated a propensity score for the likelihood of full vaccination (41), and successfully matched all 76 vaccinated to unvaccinated patients, including age, sex, county of residence, and study month (February, March, and April 2021) as covariates. The standardized mean difference between the matched pairs was 0.0263, reduced by 96.9% from 0.738 prior to matching.

**Supplementary Table 1** shows the distribution of variants in the matched pairs. McNemar’s test of the null hypothesis of equal distributions between vaccinated and unvaccinated patients, assessing the three VOC/VOI separately, could not be calculated due to sparse data. When we collapsed the table to reflect all VOCs/VOIs compared to other variants, McNemar’s test resulted in a p-value of 0.692 (**Supplementary Table 2**). Thus, vaccinated and unvaccinated individuals in the metropolitan New York area were similarly affected by the regionally circulating VOCs and VOIs. In addition, there was no clear association among vaccinated patients between type of vaccine received and Pango lineage (chi-squared test, p=0.63).

### Widespread phylogenetic dispersal of vaccine breakthrough sequences among unvaccinated controls

As a way to ascertain potential bias in our sampling, we carried out a phylogenetic analysis of our 76 breakthrough sequences in the context of 1,046 unvaccinated SARS-CoV-2-positive controls (selected randomly as part of our greater New York catchment area genomic surveillance) together with sub-sampled sequences from the United States as well as globally, the latter two groups based on Nextstrain builds of GISAID sequences (**Figure 2**). The main variants circulating in the New York area (purple) between the months of February and April 2021 were B.1.1.7 and B.1.526. Accordingly, our vaccine breakthrough samples (orange branch symbols and gray rays) mostly engaged B.1.1.7 and B.1.526 clades and were interspersed among the unvaccinated controls as well as other United States sequences. There was no evidence of extensive clustering that might indicate onward transmissions or transmission chains of vaccine breakthrough infections. Instead, they were widely distributed and appeared to mostly involve independent clusters of infections.

**Figure 2.**
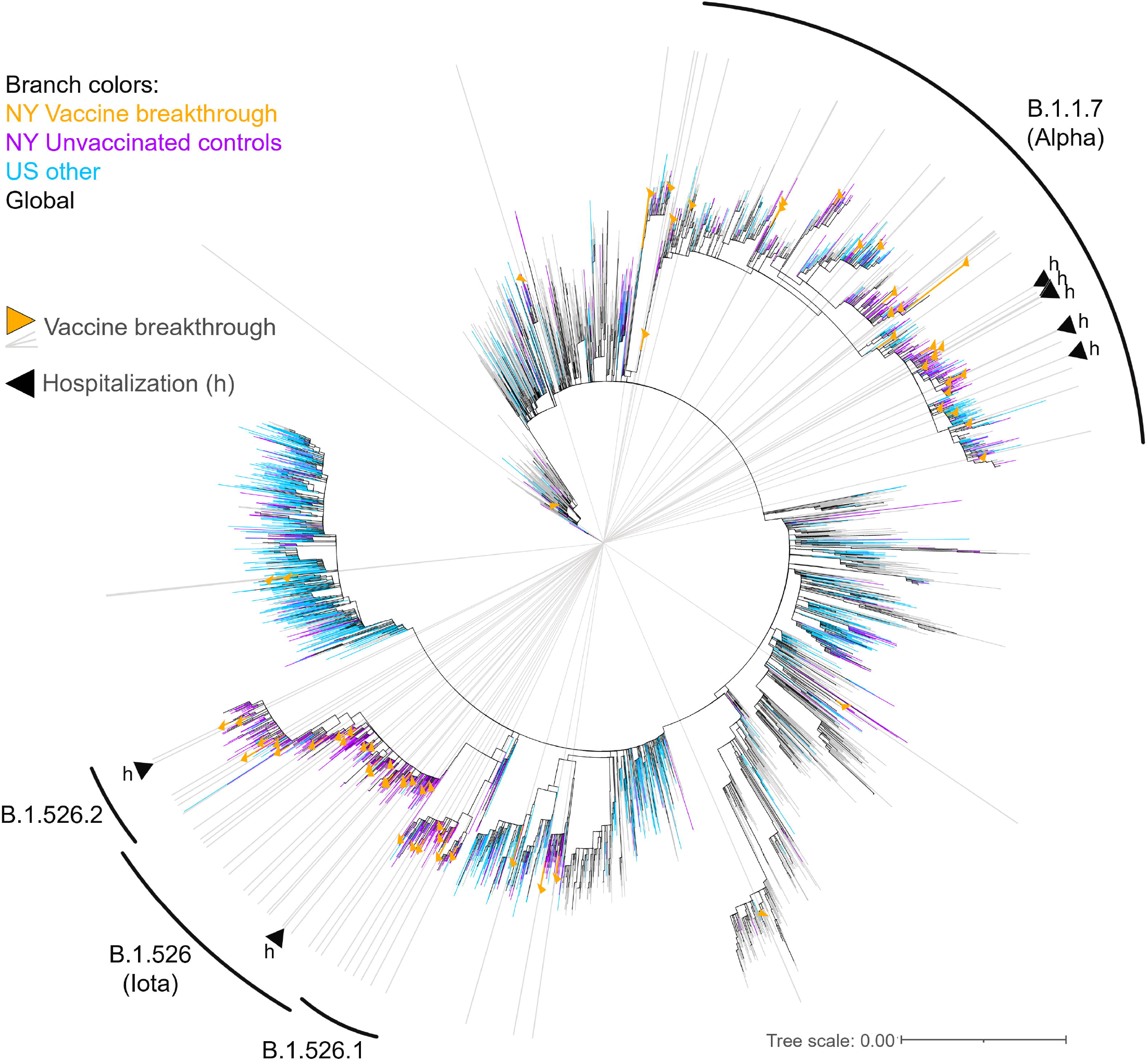
Maximum likelihood tree of SARS-CoV-2 vaccine breakthrough, unvaccinated matched control, and global reference sequences. IQ tree of 4,923 SARS-CoV-2 full genome sequences (base pairs 202-29,657 according to Wuhan-Hu-1 as reference), including 76 vaccine breakthrough (orange) and 1,046 unvaccinated control SARS-CoV-2 sequences from our NYU cohort (greater NYC area) (purple) together with 1,361 other US (cyan) and 2,440 non-US global reference sequences (black). The tree was generated with a GTR+I+G substitution model and 1,000 bootstrap replicates, and the substitution scale of the tree is indicated at the bottom right. The branches of the tree are colored as indicated. Vaccine breakthrough sequences are highlighted by orange triangles as branch symbols and gray rays radiating from the root to the outer rim of the tree. Hospitalizations due to COVID-19 among the vaccine breakthrough infections are indicated by black triangles (h). The variants responsible for most vaccine breakthrough infections in our study cohort are labeled with respective Pango lineages (WHO classification in parenthesis).

### Enrichment of NTD deletions and RBD escape mutations in vaccine breakthrough compared to unvaccinated control sequences

To screen whether vaccine breakthrough preferentially occurred with distinct vaccine escape mutations, we performed a comparative analysis of spike mutations between case and control groups. In our aligned data set of 76 vaccine breakthrough and 1,046 unvaccinated control sequences, spike mutations (compared to Wuhan-Hu-1) occurred at 230 amino acid residues. While most of these sites were more frequently mutated in control cases (182 sites), and one site (D614) being equally mutated in both groups (100% D614G), 47 sites exhibited increased mutation rates in the vaccine breakthrough group (**Figure 3A**). Interestingly, the degree of enrichment (Δ mut) was higher at the 47 breakthrough-enriched sites compared to the 182 sites enriched in controls. Contributing factors presumably included random mutations in the absence of immune pressure in controls, adaptive selection of immune escape mutations in vaccine breakthroughs, but also uneven case numbers in both groups. When we disregarded unique mutations per data set in our calculations, the mutation analysis yielded 23 distinct spike sites with enriched mutations in breakthrough infections (**Figure 3A, Supplementary Table 4**). Although individual sites did not achieve significance in Fisher’s exact tests, the array of sieved mutation sites drew a striking pattern of N-terminal domain (NTD) deletions (ΔY144 and ΔV69-H70), receptor-binding domain (RBD) mutations (E484K, N501Y, and K417N/T), an S1 mutation known to modulate the RBD up or down positioning (A570D/V) (1), a mutation right in front of the furin binding site known to affect/improve S1/S2 cleavage (P681H/R) (2), and also C-terminal mutations in S2 (T716I, S982A, T1027I, and D1118H), all of which have been associated with enhanced immune evasion, ACE2 receptor binding, and/or recurring in VOCs/VOIs (3, 4). The overrepresentation was most pronounced for E484K, followed by A570D/V, P681H/R, and ΔY144, which surpassed background spike mutation levels in unvaccinated controls (compared to Wuhan-Hu-1) by more than 12-fold. Higher levels of NTD deletion ΔY144 as well as S1 mutation A570D/V was based on the (non-significant) overrepresentation of the B.1.1.7 variant in breakthrough cases (34.2%) compared to non-vaccinated controls (29.0%), and, in the case of ΔY144, was supported by a slight difference in frequencies of sublineage B.1.526.1 with its characteristic ΔY144 deletion in breakthrough (9.2%) versus control cases (8.4%) (**Supplementary Figure 1**). Higher levels of P681H/R mutations in breakthrough cases traced back to infections with B.1.1.7 but also other viruses carrying this mutation, e.g., within lineage B.1.575 and B.1.1.519. RBD E484K mutations were found in different B.1.526 subsets (Iota and B.1.526.2 sublineage) and occurred more frequently in breakthrough compared to control cases (**Supplementary Figure 1**).

**Figure 3.**
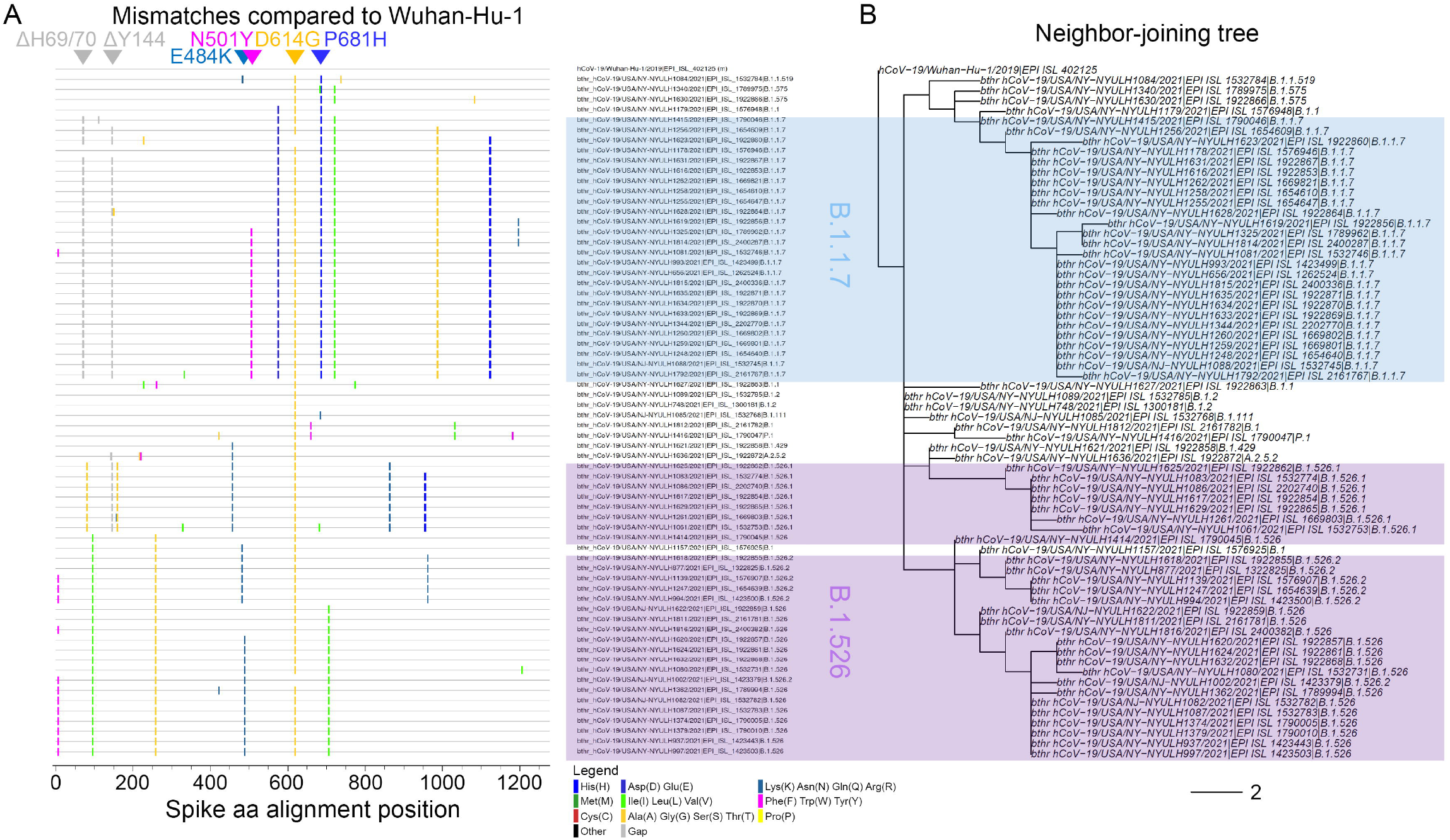
Site-specific spike mutation analysis in SARS-CoV-2 vaccine breakthrough sequences compared to unvaccinated matched controls. **A**) Comparison of site-specific amino acid mutation (mut) frequencies in spike in 76 vaccine breakthrough sequences (Vacc) compared to 1,046 unvaccinated matched controls (Unvacc) from the same cohort. The Wuhan-Hu-1 sequence served as reference to call mutations per site, and all spike mutation sites of the study sequences are shown along the x-axis according to their spike position (n=230). The mirror plot displays differences of mutation frequencies between Vacc and Unvacc groups; orange bars (top) refer to higher mutation rates in Vacc sequences, whereas black bars (bottom) refer to higher mutation rates in Unvacc matched controls. **B**) Enrichment of spike mutations in SARS-CoV-2 vaccine breakthrough sequences. All sites with greater spike mutation rates in Vacc compared to Unvacc controls are shown; sites with unique occurrences of mutations in breakthrough cases were disregarded. Mutation sites in the spike N-terminal domain (NTD), receptor binding domain (RBD), the C-terminal S1 region affecting RBD, and S1/S2 interface region that have been associated with variants of concern, neutralization immune escape, and/or affecting important biological functions of the virus are highlighted. The dashed black line indicates the average mutation frequency across all spike residues in the unvaccinated control data set compared to Wuhan-Hu-1 as reference (n=1,046).

## DISCUSSION

Our data from a large metropolitan healthcare system in the greater New York City area underline a high vaccine efficacy in fully vaccinated individuals, more than 14 days post-last dose of vaccination. Efficacy is maintained against several circulating variants including VOCs and VOIs. Compared to the large number of SARS-CoV-2 infections among unvaccinated individuals, the recorded breakthrough cases between February and April 2021 (n=76) remained at ∼1% of total infections, with the caveat that both breakthrough cases as well as unvaccinated controls were not exhaustively screened and covered.

Despite the overall effectiveness of vaccination, our full spike mutation analysis revealed a broad set of spike mutations (n=23) to be elevated in the vaccine breakthrough group. It indicates that adaptive selection is in progress that may subsequently come into full effect. At this point, the breakthrough cases and differences in mutation rates between case and control groups are still too low to draw meaningful conclusions. However, the modest overrepresentation of functionally important spike mutations including NTD deletions ΔH69-V70 and ΔY144 together with RBD mutations E484K and N501Y as well as A570D/V (S1 mutation modulating RBD up/down positioning) and P681H/R (next to the S1/S2 interface tuning cleavage) may indicate a starting sieve effect at individual or combinations of functional mutations. Spike mutations and deletions reported to confer neutralization escape *in vitro* (5-7) or regulating biological processes of the virus (1, 2) might thus be responsible for a sieve effect in a real-life situation, i.e., among vaccinated individuals.

During the time of our sample and data collection, there were two major variants arising in the New York City metro area constituting about two thirds of all cases, B.1.1.7, which was first reported in the UK (42), and B.1.526, which arose in New York City around December of 2020 (32, 33). B.1.1.7 was deemed a variant of concern by the WHO and CDC and was associated with higher viral loads in infected individuals (39, 40), enhanced epidemiological spread (42-44), an extended window of acute infection (45), less effective innate and adaptive immune clearance (46), and increased death rates in elderly patients and/or individuals with comorbidities (47-50).

The B.1.1.7 variant, e.g., through the RBD N501Y mutation but also through NTD deletions, acquired an enhanced affinity to ACE2, higher infectivity and virulence (51-54), while maintaining sensitivity to neutralization though with slightly impaired nAb titers (51, 55-57). The B.1.526 variant and its derivatives were designated VOI because of the presence of RBD mutations such as E484K or S477N that favor immune evasion (6, 33).

To study whether the more transmissible B.1.1.7 or B.1.526 because of its critical RBD mutations and prevalence/place of origin in our city were overrepresented in the breakthrough cases, we performed a comparative statistical analysis. Extending a few other studies reported so far (8, 22-29), we focused on a strict threshold of > 14 days post last dose of vaccination, and we performed both unmatched and matched analyses side-by-side. We adjusted for the confounding factors of sex, age, study month, and residence area, though we are aware that other confounding factors resulting from differences in sampling or behavioral factors between groups might also play a role. Therefore, we cannot guarantee a representative set of breakthrough infections; it is possible that infected individuals post-vaccination had milder symptoms and were less likely to seek testing, narrowing down the pool of breakthrough infections to more severe cases including VOCs/VOIs.

These caveats reinforce our finding that B.1.1.7 and B.1.526 are not preferentially represented in the vaccine breakthrough group but distributed at similar proportions as other variants in case and control groups, implying that the studied mRNA vaccines are comparably effective against these predominant variants in the NYC area. This is in agreement with recent data from Israel evaluating B.1.1.7 in this context (8). A sieve effect of the B.1.526 variant has not been studied in this detail, except for two studies with small sample sizes of n=2 and n=11 that did not allow strong conclusions (23, 27). A recent CDC study reported a total number of 10,262 SARS-CoV-2 vaccine breakthrough infections across the USA as of end-April 2021 causing hospitalization in 10% of cases (n=995) (28), a rate identical to our study.

Interestingly, the median Ct of our breakthrough infections, including those yielding an inadequate genome, was 26, with 50% of them (51 samples) exhibiting Ct values ≤ 25. It implies a moderate to high viral load in many of our breakthrough cases, at least in the nasopharynx from where our samples were collected. These moderate to high viral loads suggest the possibility of transmission to others high viral loads suggest the possibility of transmission to others (58, 59). Although our phylogenetic analysis does not provide evidence of transmission to others from our breakthrough cases at this time, this should be expected with the growing number of breakthrough cases in the next months. This may have significant epidemiologic and clinical significance if transmissible breakthrough infections carry specific spike mutations associated with immune evasion.

In conclusion, our data indicate that vaccine breakthrough in fully mRNA-vaccinated individuals is not more frequent with the VOC Alpha or the NY local VOI Iota, which underlines the high efficacy of mRNA COVID-19 vaccines against different variants. The increased presence of mutations in key regions of the spike protein in the vaccine breakthrough group is of potential concern and requires continued monitoring. Genomic surveillance of vaccine breakthrough cases should be carried out on a broader scale throughout the United States and the entire world to achieve higher case numbers and the inclusion of different VOCs and VOIs.

## METHODS

### Study design and sample collection

The design is an observational case-control study of SARS-CoV-2 vaccine breakthrough infections in the NYU Langone Health system, a large healthcare system in the New York City metro area, with primary care hospitals located in Manhattan (New York County), Brooklyn (Kings County) and Nassau County (Long Island). The case group consisted of individuals who tested positive by real-time RT-PCR for SARS-CoV-2 RNA regardless of Ct, any time after 14 days of inoculation with the second dose of BNT162b2 (Pfizer/BioNTech) or mRNA-1273 (Moderna) vaccines, or with the single dose COVID Janssen vaccine, according to our electronic health records (EHR). The control group consisted of full-genome sequenced SARS-CoV-2 positive cases in our health system who were randomly selected for SARS-CoV-2 genomic surveillance, had Ct ≤30, and were collected in the same time period as the breakthrough infections. Nasopharyngeal swabs were sampled from individuals suspected to have an infection with SARS-CoV-2 as part of clinical diagnostics or hospital admittance. Samples were collected in 3 mL viral transport medium (VTM; Copan universal transport medium or equivalent). Clinical testing was performed using various FDA emergency use authorization (EUA)–approved platforms for detection of SARS-CoV-2, i.e., the Roche Cobas 6800 SARS-CoV-2 (90% of the samples in this study) and Cepheid Xpert SARS-CoV-2 or SARS-CoV-2/Flu/RSV assays.

### RNA extraction, cDNA synthesis, library preparation and sequencing

RNA was extracted from 400 μl of each nasopharyngeal swab specimens using the MagMAX™ Viral/Pathogen Nucleic Acid Isolation Kit on the KingFisher flex system (Thermo Fisher Scientific) following the manufacturer’s instructions. Total RNA (11 μl) was converted to first strand cDNA by random priming using the Superscript IV first-strand synthesis system (Invitrogen, ref# 180901050). Libraries were prepared using Swift Normalase Amplicon SARS-CoV-2 Panel (SNAP) and SARS-CoV-2 additional Genome Coverage Panel (Cat# SN-5X296 core kit, 96rxn), using 10 μl of first strand cDNA, following the manufacturer’s instructions (60). Final libraries were run on Agilent Tapestation 2200 with high sensitivity DNA Screentape to verify the amplicon size of about 450 bp. Normalized pools were run on the Illumina NovaSeq 6000 system with the SP 300 cycle flow cell. Run metrics were paired-end 150 cycles with dual indexing reads. Typically, two pools representing two full 96 well plates (192 samples) were sequenced on each SP300 NovaSeq flow cell.

### Sequenced read processing

Sequencing reads were demultiplexed using the Illumina bcl2fastq2 Conversion Software v2.20 and adapters and low-quality bases were trimmed with Trimmomatic v0.36 (61). BWA v0.7.17 (62) was utilized for mapping reads to the SARS-CoV-2 reference genome (NC_045512.2, wuhCor1) and mapped reads were soft-clipped to remove SNAP tiled primer sequences using Primerclip v0.3.8 (63). BCFtools v1.9 (64) was used to call mutations and assemble consensus sequences, which were then assigned phylogenetic lineage designations according to PANGO nomenclature (65). Sequences that did not yield a near-complete viral genome (<23,000bp, <4000x coverage) were discarded from further analysis. All sequences are publicly available in SRA (BioProject PRJNA751078) and deposited in GISAID (GISAID accession numbers are in **Supplemental Table 3**).

### Phylogenetic analyses

SARS-CoV-2 full genome sequences were aligned using Mafft v.7 (66). The alignment was cropped to base pairs 202-29657 according to the Wuhan-Hu-1 reference to remove N- and C-terminal regions with unassigned base pairs. Maximum likelihood IQ trees were performed using the IQ-TREE XSEDE tool, multicore version 2.1.2, on the Cipres Science gateway v.3.3 (67). GTR+I+G was chosen as the best-fit substitution model. Support values were generated with 1,000 bootstrap replicates and the ultrafast bootstrapping method. Phylogenetic trees were visualized in Interactive Tree Of Life (iTOL) v.6 (68). The tree was constructed using 76 vaccine breakthrough and 1,046 unvaccinated control SARS-CoV-2 sequences from our NYU cohort (greater NYC area) together with 3,801 global reference sequences for a total of 4,923 SARS-CoV-2 genomic sequences. The reference sequences were retrieved from a Nextstrain build with North America-focused global subsampling [4] and included 1,361 other US sequences.

### Mutation analysis

Spike amino acid counts and calculations of site-specific mutation frequencies compared to Wuhan-Hu-1 as reference were done on MAFFT-aligned SARS-CoV-2 sequences using R(69) and R Studio(70) using scripts based on the seqinr and tidyverse (dplyr, lubridate, magrittr) packages. The calculations exclusively considered residues that were accurately covered by sequencing, i.e., non-ambiguous characters and gaps. Fisher exact tests and multiplicity corrections (Benjamini-Hochberg) were done in Program R/RStudio. Multiplicity corrected P values (q) <0.05 were considered significant. Highlighter analyses were performed on MAFFT-aligned SARS-CoV-2 amino acid sequences of the spike genomic region. SARS-CoV-2 vaccine breakthrough sequences were compared to Wuhan-Hu-1 as master using the Highlighter tool provided by the Los Alamos HIV sequence database (71).

### Statistical analysis

To address confounding and other sources of bias arising from the use of observational data, we estimated a propensity score for the likelihood of full vaccination, and matched vaccinated to unvaccinated patients, including age, sex, county of residence, and study month (February, March, April 2021) as covariates (41). Propensity-score matching was implemented using the nearest neighbor strategy using a 1:1 ratio without replacement, with the MatchIt algorithm in RStudio version 1.4.1106 (72). Before and after matching, we evaluated the presence of three Pango lineages, B.1.1.7, B.1.526 and P.1 compared to all others lineages combined. On unmatched data, we calculated a Pearson chi-squared test statistic; on matched data we calculated McNemar’s test. The tables used for these calculations are in Supplementary Material (**Supplementary Tables 1 and 2**). Comparisons of Ct values and mutation counts between groups were made using non-parametric Mann-Whitney tests or Kruskal-Wallis tests with Dunn’s multiplicity correction.

### Study Approval

This study was approved by the NYU Langone Health Institutional Review Board, protocol number i21-00493.

## Supporting information

Supplemental Table 5-GISAID acknowledgements

## Data Availability

The SARS-C0V-2 sequences have been uploaded to GISAID

## Author contributions

Ralf Duerr, MD, PhD designed the study, analyzed data and wrote the manuscript; Dacia Dimartino, PhD generated sequencing data; Christian Marier analyzed genomic data; Paul Zappile generated sequencing data; Guiqing Wang, MD, PhD collected samples and performed clinical SARS-CoV-2 detection; Jennifer Lighter, MD reviewed clinical information; Brian Elbel, PhD monitored epidemiological data and generated lists of breakthrough infections; Andrea Troxel, PhD performed statistical analyses; Adriana Heguy, PhD designed the study, generated genomic data and wrote the manuscript. All authors reviewed and edited the manuscript.

## Acknowledgements

The authors wish to thank Vincent Setang and NYU Langone Health DataCore for support extracting the data for this study from clinical databases, Dr. Maria Agüero-Rosenfeld and the clinical laboratory technicians for assistance in testing, saving and retrieving specimens, especially Joanna Fung for her assistance with RNA extraction project. We also thank Dr. Joan Cangiarella for her continuous support of genomic surveillance for SARS-CoV-2 at NYULH, including providing institutional funding for this study. We are grateful to the authors and submitting laboratories who deposited data in GISAID, in particular to those whose sequences we used to create the phylogenetic tree. These are all named in Supplemental Table 5. The NYULH Genome Technology Center is partially supported by the Cancer Center Support Grant P30CA016087 at the Laura and Isaac Perlmutter Cancer Center.

---

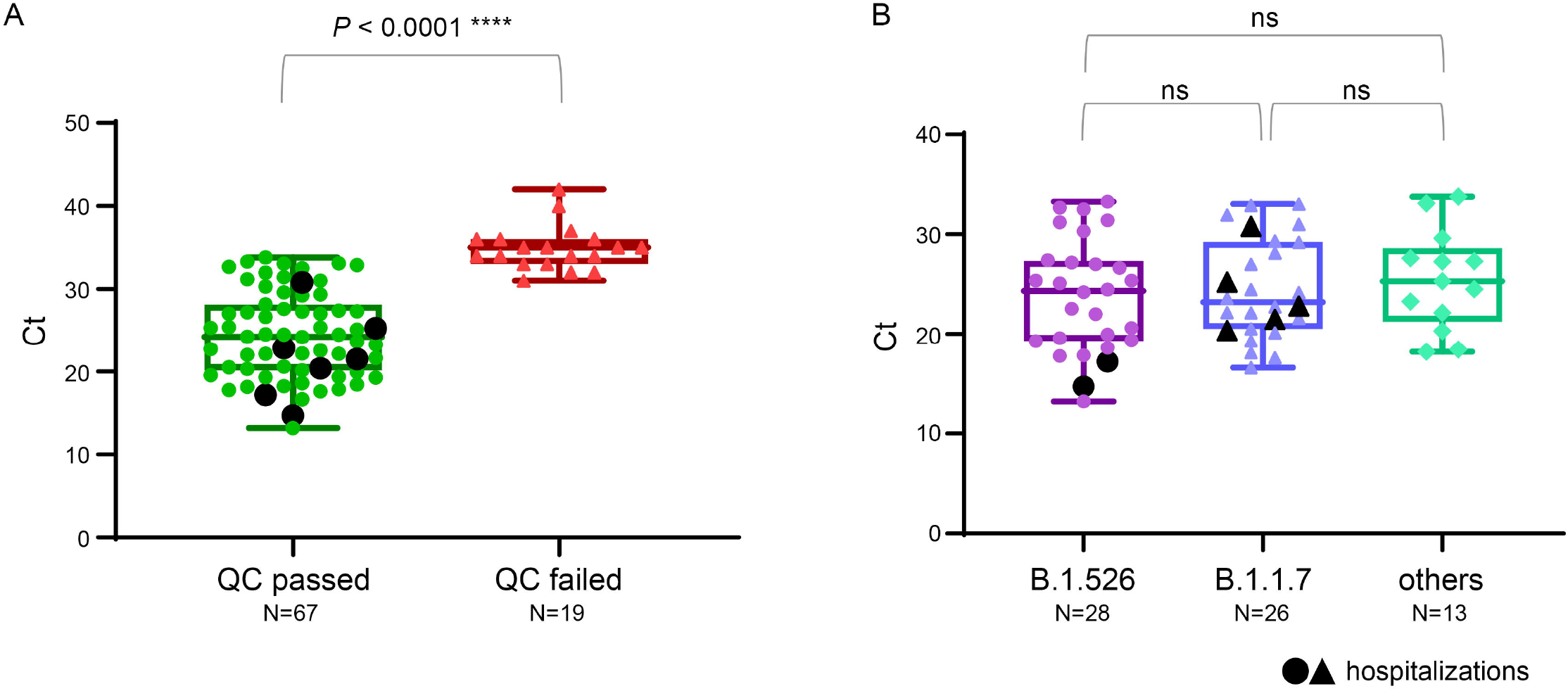

