## Supplementary material for "Dominance of Alpha and Iota variants in SARS-CoV-2 vaccine breakthrough infections in New York City": Supplememtal methods and data

**Supplementary figure legends.**

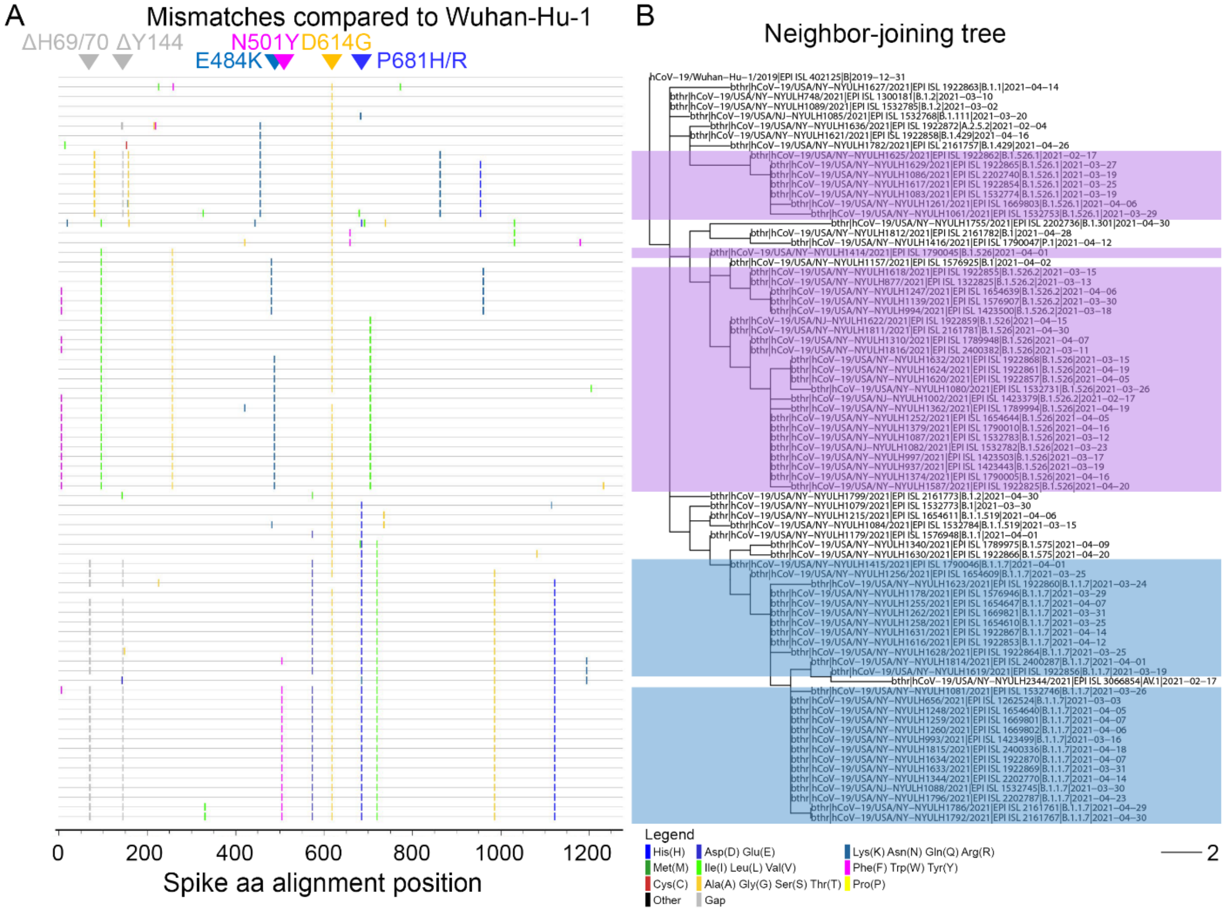


**Supplementary Figure 1**. Spike mutation patterns in vaccine breakthrough sequences. **A**) Highlighter plot showing spike amino acid mutations of 76 vaccine breakthrough SARS-CoV-2 sequences compared to the Wuhan-Hu-1 reference sequence as master (top line). Mutations are shown as ticks, color-coded according to the legend at the bottom. Key mutations are indicated by triangles and labeled. Study sequences are sorted according to the Neighbor-joining tree to the right (**B**). The scale bar at the bottom indicates the branch length of two substitutions. B.1.1.7 and B.1.526 sequences are highlighted in blue and purple, respectively.

**Supplementary Tables.**

Supplementary Table 1: Numbers of SARS-CoV-2 breakthrough and unvaccinated control infections in matched pairs in New York according to Pango lineages.

| | | Vaccinated | | | | |
|---|---|---|---|---|---|---|
| | | B.1.1.7 | B.1.526 | P.1 | other | Total |
| Unvaccinated | B.1.1.7 | 10 | 11 | 0 | 8 | 29 |
| | B.1.526 | 10 | 10 | 1 | 5 | 26 |
| | P.1 | 1 | 1 | 0 | 1 | 3 |
| | other | 5 | 9 | 0 | 4 | 18 |
| | Total | 26 | 31 | 1 | 18 | 76 |

Supplementary Table 2: Numbers of SARS-CoV-2 breakthrough and unvaccinated control infections in matched pairs in New York according to VOC/VOI or non-VOC/VOI.

| | | Vaccinated | | |
|---|---|---|---|---|
| | | VOC/VOI | other | Total |
| Unvaccinated | VOC/VOI | 46 | 11 | 57 |
| | other | 9 | 1 | 10 |
| | Total | 55 | 12 | 67 |

### Supplementary Table 3: Full specifications of breakthrough infections

| Sample ID | Days after completion of vaccination series | Vaccine | Hospitalization | Sex | Age Range | CT | State | County | GISAID Accession | GISAID Virus Name | GISAID Clade | Pango Lineage | AA Substitutions |
|---|---|---|---|---|---|---|---|---|---|---|---|---|---|
| P21-0524 | 28 | Pfizer | N | M | 21-30 | 33.3 | NJ | Bergen | EPI_ISL_1423379 | NYULH1002 | GH | B.1.526.2 | Spike A701V, Spike D253G, Spike E484K, Spike L5F, Spike T95I, N M234I, NS3 P42L, NS3 Q57H, NS8 T11I, NSP2 T85I, NSP4 L438P, NSP6 F108del, NSP6 G107del, NSP6 S106del, NSP12 P323L, NSP13 Q88H |
| P21-0852 | 54 | Pfizer | N | F | 21-30 | 22.2 | NY | New York | EPI_ISL_1262524 | NYULH656 | GRY | B.1.1.7 | Spike A570D, Spike D614G, Spike D1118H, Spike H69del, Spike N501Y, Spike P681H, Spike S982A, Spike T716I, Spike V70del, Spike Y144del, M G89S, N A398V, N D3L, N G204R, N R203K, N S235F, NS3 L15F, NS8 Q27stop, NS8 R52I, NS8 Y73C, NSP3 A890D, NSP3 I1412T, NSP3 T183I, NSP4 F17L, NSP6 F108del, NSP6 G107del, NSP6 S106del, NSP12 P323L, NSP13 K460R, NSP14 S171G |
| P21-1038 | 22 | Moderna | N | F | 71-80 | 27.3 | NY | Suffolk | EPI_ISL_1300181 | NYULH748 | GH | B.1.2 | Spike D614G, N D377Y, N P67S, N P199L, NS3 G172V, NS3 Q57H, NS8 S24L, NSP2 T85I, NSP3 D59G, NSP3 M829I, NSP4 M458I, NSP5 L89F, NSP8 A14V, NSP12 K59R, NSP12 P323L, NSP14 N129D, NSP16 R216C |
| P21-1234 | 21 | Pfizer | N | M | 71-80 | 25.4 | NY | Nassau | EPI_ISL_1423443 | NYULH937 | GH | B.1.526 | Spike A701V, Spike D253G, Spike D614G, Spike E484K, Spike L5F, Spike T95I, N M234I, N P199L, NS3 P42L, NS3 Q57H, NS8 T11I, NSP1 H110Y, NSP2 T85I, NSP4 L438P, NSP6 F108del, NSP6 G107del, NSP6 S106del, NSP12 P323L, NSP13 Q88H |
| P21-1292 | 33 | Pfizer | Y (COVID) | F | 71-80 | 20.4 | NY | Kings | EPI_ISL_1423499 | NYULH993 | GRY | B.1.1.7 | Spike A570D, Spike D614G, Spike D1118H, Spike H69del, Spike N501Y, Spike P681H, Spike S982A, Spike T716I, Spike V70del, Spike Y144del, N D3L, N G204R, N R203K, N S235F, NS8 K68stop, NS8 Q27stop, NS8 R52I, NS8 Y73C, NSP3 A890D, NSP3 I1412T, NSP3 M1441I, NSP3 T183I, NSP5 D34G, NSP6 F108del, NSP6 G107del, NSP6 S106del, NSP12 M519L, NSP12 P227L, NSP12 P323L, NSP13 A509V |
| P21-1293 | 58 | Pfizer | N | F | 31-40 | 27.4 | NY | New York | EPI_ISL_1423500 | NYULH994 | GH | B.1.526.2 | Spike D253G, Spike D614G, Spike L5F, Spike Q957R, Spike S477N, Spike T95I, N P13L, N S202R, NS3 P42L, NS3 Q57H, NS7a L116F, NS7a R78C, NS8 T11I, NSP2 T85I, NSP3 G1128S, NSP4 L438P, NSP6 F108del, NSP6 G107del, NSP6 S106del, NSP12 P323L, NSP13 Q88H |
| P21-1296 | 40 | Pfizer | N | F | 41-50 | 31.4 | NY | New York | EPI_ISL_1423503 | NYULH997 | GH | B.1.526 | Spike A701V, Spike D253G, Spike D614G, Spike E484K, Spike L5F, Spike T95I, N H300Y, N I94V, N M234I, N P199L, NS3 P42L, NS3 Q57H, NS8 T11I, NSP2 T85I, NSP4 L438P, NSP6 F108del, NSP6 G107del, NSP6 S106del, NSP12 P323L, NSP13 Q88H |
| P21-1362 | 61 | Pfizer | N | F | 41-50 | 19.9 | NY | New York | EPI_ISL_1532753 | NYULH1061 | GH | B.1.526.1 | Spike D80G, Spike D614G, Spike D950H, Spike F157S, Spike L452R, Spike T323I, Spike T676I, Spike T859N, Spike Y144del, N M234I, N T205I, NS3 P42L, NS3 P104L, NS3 Q57H, NS8 A51S, NS8 T11I, NSP2 T85I, NSP3 A861S, NSP4 A446V, NSP4 L438P, NSP6 F108del, NSP6 G107del, NSP6 S106del, NSP6 V278I, NSP12 P323L, NSP16 K160R |
| P21-1394 | 70 | Pfizer | N | F | 21-30 | 32.0 | NY | Rockland | EPI_ISL_1654609 | NYULH1256 | GR | B.1.1.7 | Spike A570D, Spike D614G, Spike H69del, Spike P681H, Spike S982A, Spike T716I, Spike V70del, Spike Y144del, N D3L, N G204R, N R203K, N S235F, NS8 K68stop, NS8 Q27stop, NS8 R52I, NS8 Y73C, NSP6 R138del, NSP6 V139del, NSP6 W140del, NSP13 A509V, NSP13 P234L |
| P21-1396 | 70 | Pfizer | N | F | 31-40 | 27.0 | NY | Nassau | EPI_ISL_1654610 | NYULH1258 | GR | B.1.1.7 | Spike A570D, Spike D614G, Spike D1118H, Spike H69del, Spike P681H, Spike S982A, Spike T716I, Spike V70del, Spike Y144del, E L21F, N D3L, N G204R, N R203K, N S235F, NS7a I103del, NS8 Q27stop, NS8 R52I, NS8 Y73C, NSP2 K489E, NSP3 A890D, NSP3 I1412T, NSP3 T183I, NSP6 F108del, NSP6 G107del, NSP6 S106del, NSP12 A399T, NSP12 P323L, NSP16 S241P |
| P21-1402 | 20 | Janssen | Y (COVID) | F | 51-60 | 25.0 | NY | Kings | EPI_ISL_1532773 | NYULH1079 | GH | B.1 | Spike D614G, Spike E1111K, Spike P681H, N M234I, NS3 Q57H, NSP2 T85I, NSP4 T189I, NSP4 T439M, NSP6 H11Q, NSP9 P57S, NSP12 P323L |
| P21-1403 | 72 | Pfizer | N | F | 21-30 | 27.2 | NY | Kings | EPI_ISL_1532731 | NYULH1080 | GH | B.1.526 | Spike A701V, Spike D253G, Spike D614G, Spike E484K, Spike Q1201L, Spike T95I, N M234I, N P199L, NS3 A33S, NS3 P42L, NS3 Q57H, NS8 T11I, NSP2 T85I, NSP3 T1189I, NSP4 L438P, NSP4 S218F, NSP6 F108del, NSP6 G107del, NSP6 S106del, NSP12 P323L, NSP13 Q88H, NSP16 L163F |
| P21-1404 | 73 | Pfizer | N | F | 41-50 | 16.6 | NY | Richmond | EPI_ISL_1532746 | NYULH1081 | GRY | B.1.1.7 | Spike A570D, Spike D614G, Spike D1118H, Spike H69del, Spike L5F, Spike N501Y, Spike P681H, Spike S982A, Spike T716I, Spike V70del, Spike Y144del, N D3L, N G204R, N R203K, N S235F, NS3 T151I, NS7a Q62stop, NS8 Q27stop, NS8 R52I, NS8 Y73C, NSP2 L137F, NSP3 A890D, NSP3 I1412T, NSP3 P340L, NSP3 T183I, NSP3 T1303I, NSP6 F108del, NSP6 G107del, NSP6 S106del, NSP12 P323L, NSP12 T739I |
| P21-1405 | 58 | Pfizer | N | F | 31-40 | 25.1 | NJ | Essex | EPI_ISL_1532782 | NYULH1082 | GH | B.1.526 | Spike A701V, Spike D253G, Spike D614G, Spike E484K, Spike L5F, Spike T95I, N M234I, N P199L, NS3 P42L, NS3 Q57H, NS8 T11I, NSP2 T85I, NSP4 L438P, NSP6 F108del, NSP6 G107del, NSP6 S106del, NSP12 P323L, NSP13 Q88H, NSP14 T428A |
| P21-1406 | 65 | Pfizer | N | F | 31-40 | 13.2 | NY | Bronx | EPI_ISL_1532774 | NYULH1083 | GH | B.1.526.1 | Spike D80G, Spike D614G, Spike D950H, Spike F157S, Spike L452R, Spike T859N, Spike Y144del, N M234I, N T205I, NS3 P42L, NS3 P104L, NS3 Q57H, NS8 A51S, NS8 T11I, NSP2 T85I, NSP4 A446V, NSP4 K399E, NSP4 L438P, NSP6 F108del, NSP6 G107del, NSP6 S106del, NSP6 V278I, NSP12 P323L |
| P21-1407 | 49 | Pfizer | N | M | 31-40 | 27.6 | NY | New York | EPI_ISL_1532784 | NYULH1084 | GR | B.1.1.519 | Spike D614G, Spike P681H, Spike T478K, Spike T732A, N G204R, N R203K, NSP1 G112C, NSP3 P141S, NSP3 V1704F, NSP4 T492I, NSP6 I49V, NSP6 L37F, NSP9 T35I, NSP12 P323L, NSP15 L162F |
| P21-1411 | 54 | Pfizer | N | F | 51-60 | 29.6 | NJ | Bergen | EPI_ISL_1532768 | NYULH1085 | GH | B.1.111 | Spike D614G, Spike N679K, N K387R, N T205I, NS3 G44E, NS3 Q57H, NS3 T223I, NS8 L57S, NSP1 I114T, NSP12 P323L |
| P21-1412 | 62 | Pfizer | N | F | 51-60 | 29.2 | NY | Queens | EPI_ISL_2202740 | NYULH1086 | GH | B.1.526.1 | Spike D80G, Spike D614G, Spike D950H, Spike F157S, Spike L452R, Spike T859N, Spike Y144del, N E323K, N M234I, N T205I, NS3 P42L, NS3 P104L, NS3 Q57H, NS8 A51S, NS8 T11I, NSP2 T85I, NSP3 A861S, NSP4 A446V, NSP4 K399E, NSP4 L438P, NSP6 F108del, NSP6 G107del, NSP6 S106del, NSP6 V278I, NSP12 P323L, NSP13 A469T |
| P21-1413 | 67 | Pfizer | N | M | 31-40 | 17.2 | NY | Nassau | EPI_ISL_1532783 | NYULH1087 | GH | B.1.526 | Spike A701V, Spike D253G, Spike D614G, Spike E484K, Spike L5F, Spike T95I, N M234I, N P199L, NS3 P42L, NS3 P104L, NS3 Q57H, NS8 T11I, NSP2 T85I, NSP4 L438P, NSP6 F108del, NSP6 G107del, NSP6 S106del, NSP12 P323L, NSP13 Q88H, NSP14 A1S, NSP16 P236L |
| P21-1415 | 42 | Moderna | N | M | 61-70 | 23.6 | NJ | Essex | EPI_ISL_1532745 | NYULH1088 | GRY | B.1.1.7 | Spike A570D, Spike D614G, Spike D1118H, Spike H69del, Spike N501Y, Spike P681H, Spike S982A, Spike T716I, Spike V70del, Spike Y144del, N D3L, N G204R, N R203K, N S235F, NS3 P240S, NS8 K68stop, NS8 Q27stop, NS8 R52I, NS8 Y73C, NSP3 A890D, NSP3 E95D, NSP3 I1412T, NSP3 M1441I, NSP3 T183I, NSP6 F108del, NSP6 G107del, NSP6 S106del, NSP12 P227L, NSP12 P323L, NSP13 T588I |

| | | | | | | | | | | | | | |
|---|---|---|---|---|---|---|---|---|---|---|---|---|---|
| P21-1416 | 45 | Pfizer | N | F | 41-50 | 27.3 | NY | Queens | EPI_ISL_1532785 | NYULH1089 | GH | B.1.2 | Spike D614G, N P13L, N P67S, N P199L, NS3 G172V, NS3 Q57H, NS8 S24L, NSP2 T85I, NSP2 T547I, NSP3 P968L, NSP5 L89F, NSP6 T181I, NSP12 P323L, NSP12 V354L, NSP13 Q275L, NSP14 N129D, NSP16 R216C |
| P21-1556 | 85 | Pfizer | N | M | 41-50 | 18.6 | NY | New York | EPI_ISL_1576907 | NYULH1139 | GH | B.1.526.2 | Spike D253G, Spike D614G, Spike L5F, Spike Q957R, Spike S477N, Spike T95I, N A414V, N P13L, N S202R, NS3 P42L, NS3 Q57H, NS7a L116F, NS8 T11I, NSP2 T85I, NSP3 A1179T, NSP3 G1128S, NSP3 T1022I, NSP4 L438P, NSP4 T319I, NSP6 F108del, NSP6 G107del, NSP6 S106del, NSP12 P323L, NSP13 Q88H |
| P21-1574 | 22 | Pfizer | N | F | 21-30 | 23.3 | NY | Suffolk | EPI_ISL_1576925 | NYULH1157 | G | B.1 | Spike D253G, Spike D614G, Spike S477N, Spike T95I, N G204R, N R203K, N S235F, NS3 P42L, NS3 Q57H, NS8 T11I, NSP2 T85I, NSP3 G1128S, NSP4 L438P, NSP6 F108del, NSP6 G107del, NSP6 S106del, NSP12 P323L, NSP13 Q88H |
| P21-1595 | 53 | Pfizer | N | F | 71-80 | 17.6 | NY | New York | EPI_ISL_1576946 | NYULH1178 | GR | B.1.1.7 | Spike A570D, Spike D614G, Spike D1118H, Spike P681H, Spike S982A, Spike T716I, N D3L, N G204R, N R203K, N S235F, NS8 T11I, NSP2 L550F, NSP3 A890D, NSP3 I1412T, NSP6 F108del, NSP6 G107del, NSP6 S106del, NSP12 P323L |
| P21-1596 | 57 | Moderna | N | M | 71-80 | 33.8 | NY | Richmond | EPI_ISL_1576948 | NYULH1179 | GR | B.1.1 | Spike A570D, Spike D614G, Spike P681H, N D3L, N G204R, N R203K, NS8 A51S, NS8 T11I, NSP6 F108del, NSP6 G107del, NSP6 S106del |
| P21-1948 | 18 | Pfizer | Y (not COVID) | F | 81-90 | 14.2 | NY | Queens | EPI_ISL_1654611 | NYULH1215 | GR | B.1.1.519 | Spike D614G, Spike P681H, Spike T732A, N G204R, N R203K, NS3 L15del, NS3 T14del, NS8 A65V, NS8 L4P, NSP3 P141S, NSP4 T492I, NSP6 I49V, NSP9 T35I, NSP12 P323L |
| P21-1964 | 78 | Pfizer | N | M | 51-60 | 20.5 | NY | Queens | EPI_ISL_1669801 | NYULH1259 | GRY | B.1.1.7 | Spike A570D, Spike D614G, Spike D1118H, Spike H69del, Spike N501Y, Spike P681H, Spike S982A, Spike T716I, Spike V70del, Spike Y144del, M G89S, N A398V, N D3L, N G204R, N R203K, N S235F, NS3 L15F, NS8 Q27stop, NS8 R52I, NS8 Y73C, NSP3 A890D, NSP3 I1412T, NSP3 T183I, NSP4 F17L, NSP6 F108del, NSP6 G107del, NSP6 S106del, NSP8 Q73K, NSP12 P323L, NSP13 K460R |
| P21-1970 | 71 | Moderna | N | F | 31-40 | 27.0 | NY | Suffolk | EPI_ISL_1669802 | NYULH1260 | GRY | B.1.1.7 | Spike A570D, Spike D614G, Spike D1118H, Spike H69del, Spike N501Y, Spike P681H, Spike S982A, Spike T716I, Spike V70del, Spike Y144del, N D3L, N G204R, N R203K, N S235F, NS3 A99T, NS3 V90F, NS7b T40I, NS8 Q27stop, NS8 R52I, NS8 Y73C, NSP3 A890D, NSP3 I1412T, NSP3 T183I, NSP6 F108del, NSP6 G107del, NSP6 S106del, NSP12 P323L, NSP13 K460R, NSP14 E347G |
| P21-1976 | 89 | Pfizer | N | M | 31-40 | 21.6 | NY | Suffolk | EPI_ISL_1669803 | NYULH1261 | GH | B.1.526.1 | Spike D80G, Spike D614G, Spike D950H, Spike F157S, Spike L452R, Spike S155R, Spike T859N, Spike Y144del, N M234I, N T205I, NS3 P42L, NS3 P104L, NS3 Q57H, NS8 A51S, NS8 T11I, NSP2 T85I, NSP3 A861S, NSP4 A446V, NSP4 K399E, NSP4 L438P, NSP6 F108del, NSP6 G107del, NSP6 S106del, NSP6 V278I, NSP12 P323L, NSP12 T806I, NSP13 T239I |
| P21-1984 | 76 | Pfizer | N | M | 51-60 | 19.4 | NY | Queens | EPI_ISL_1654639 | NYULH1247 | GH | B.1.526.2 | Spike D253G, Spike D614G, Spike L5F, Spike Q957R, Spike S477N, Spike T95I, N P13L, N S202R, NS3 P42L, NS3 Q57H, NS7a L116F, NS7b H37L, NS8 T11I, NSP2 T85I, NSP2 T434I, NSP3 G1128S, NSP4 L438P, NSP6 F108del, NSP6 G107del, NSP6 S106del, NSP12 P323L, NSP13 Q88H |
| P21-1985 | 76 | Pfizer | N | M | 21-30 | 22.7 | NY | Queens | EPI_ISL_1654640 | NYULH1248 | GRY | B.1.1.7 | Spike A570D, Spike D614G, Spike D1118H, Spike H69del, Spike N501Y, Spike P681H, Spike S982A, Spike T716I, Spike V70del, Spike Y144del, M G89S, N A398V, N D3L, N G204R, N R203K, N S235F, NS3 L15F, NS8 Q27stop, NS8 R52I, NS8 Y73C, NSP3 A890D, NSP3 I1412T, NSP3 T183I, NSP4 F17L, NSP6 F108del, NSP6 G107del, NSP6 S106del, NSP8 Q73K, NSP12 P323L, NSP13 K460R |
| P21-1989 | 26 | Janssen | N | M | 21-30 | 25.1 | NY | New York | EPI_ISL_1654644 | NYULH1252 | GH | B.1.526 | Spike A701V, Spike D253G, Spike D614G, Spike E484K, Spike L5F, Spike T95I, N M234I, N P199L, NS3 P42L, NS3 Q57H, NS8 T11I, NSP2 E558K, NSP2 T85I, NSP4 L438P, NSP6 F108del, NSP6 G107del, NSP6 S106del, NSP9 M101I, NSP12 P323L, NSP13 H164Y, NSP13 Q88H |
| P21-1992 | 76 | Pfizer | N | M | 41-50 | 32.9 | NY | New York | EPI_ISL_1654647 | NYULH1255 | GR | B.1.1.7 | Spike A570D, Spike D614G, Spike D1118H, Spike H69del, Spike P681H, Spike S982A, Spike T716I, Spike V70del, Spike Y144del, N G204R, N R203K, N S235F, NS8 S67F, NS8 T11I, NSP1 N178S, NSP2 L550F, NSP3 A890D, NSP3 I1412T, NSP3 T183I, NSP6 F108del, NSP6 G107del, NSP6 S106del, NSP12 P323L, NSP13 Q88H |
| P21-1993 | 50 | Moderna | Y (COVID) | M | 81-90 | 22.9 | NY | Nassau | EPI_ISL_1669821 | NYULH1262 | GR | B.1.1.7 | Spike A570D, Spike D614G, Spike D1118H, Spike H69del, Spike P681H, Spike S982A, Spike T716I, Spike V70del, Spike Y144del, N D3L, N G204R, N R203K, N S235F, NS3 P240S, NS3 T9I, NS8 K68stop, NS8 Q27stop, NS8 R52I, NS8 Y73C, NSP3 A890D, NSP3 E95D, NSP3 K963R, NSP3 M1441I, NSP3 T183I, NSP6 F108del, NSP6 G107del, NSP6 S106del, NSP12 P227L, NSP12 P323L, NSP16 A34V |
| P21-2057 | 61 | Pfizer | N | M | 71-80 | 16.5 | NY | Westchester | EPI_ISL_1789948 | NYULH1310 | GH | B.1.526 | Spike A701V, Spike D253G, Spike D614G, Spike L5F, Spike T95I, N M234I, N P199L, NS3 P42L, NS3 Q57H, NS8 T11I, NSP2 T85I, NSP4 L438P, NSP6 F108del, NSP6 G107del, NSP6 S106del, NSP12 P323L, NSP13 Q88H |
| P21-2096 | 88 | Pfizer | N | F | 31-40 | 33.1 | NY | Suffolk | EPI_ISL_1789975 | NYULH1340 | GH | B.1.575 | Spike D614G, Spike P681H, Spike T678I, Spike T716I, M I82T, N T205I, NS3 Q57H, NS7a L96F, NSP4 T492I, NSP6 T181I, NSP12 P323L, NSP13 T127I, NSP15 L162V |
| P21-2105 | 97 | Pfizer | N | M | 21-30 | 25.3 | NY | Queens | EPI_ISL_2202770 | NYULH1344 | GRY | B.1.1.7 | Spike A570D, Spike D614G, Spike D1118H, Spike H69del, Spike N501Y, Spike P681H, Spike S982A, Spike T716I, Spike V70del, Spike Y144del, N D3L, N G204R, N R14C, N R203K, N S235F, NS8 K68stop, NS8 Q27stop, NS8 R52I, NS8 Y73C, NSP3 A890D, NSP3 I1412T, NSP3 T183I, NSP6 F108del, NSP6 G107del, NSP6 P198L, NSP6 S106del, NSP12 P227L, NSP12 P323L |
| P21-2124 | 61 | Pfizer | N | F | 21-30 | 24.5 | NY | Queens | EPI_ISL_1789994 | NYULH1362 | GH | B.1.526 | Spike A701V, Spike D253G, Spike D614G, Spike E484K, Spike K417N, Spike L5F, Spike T95I, N M234I, N P199L, NS3 P42L, NS3 Q57H, NS8 T11I, NSP2 T85I, NSP4 L438P, NSP6 F108del, NSP6 G107del, NSP6 S106del, NSP12 P323L, NSP13 Q88H |
| P21-2137 | 67 | Moderna | N | F | 31-40 | 17.8 | NY | New York | EPI_ISL_1790005 | NYULH1374 | GH | B.1.526 | Spike A701V, Spike D253G, Spike D614G, Spike E484K, Spike L5F, Spike T95I, N M234I, N P199L, NS3 P42L, NS3 Q57H, NS8 A65V, NS8 T11I, NSP2 T85I, NSP4 L438P, NSP6 F108del, NSP6 G107del, NSP6 S106del, NSP12 P323L, NSP13 Q88H |
| P21-2143 | 55 | Pfizer | Y (not COVID) | F | 81-90 | 19.6 | NY | Kings | EPI_ISL_1790010 | NYULH1379 | GH | B.1.526 | Spike A701V, Spike D253G, Spike D614G, Spike E484K, Spike L5F, Spike T95I, N M234I, N P199L, NS3 P42L, NS3 Q57H, NS6 D30del, NS6 F22del, NS6 I26del, NS6 K23del, NS6 L29del, NS6 N28del, NS6 S25del, NS6 V24del, NS6 W27del, NS8 T11I, NSP2 T85I, NSP4 L438P, NSP6 F108del, NSP6 G107del, NSP6 I210T, NSP6 S106del, NSP12 P323L, NSP13 Q88H |
| P21-2191 | 68 | Pfizer | N | M | 31-40 | 17.9 | NY | Kings | EPI_ISL_1790045 | NYULH1414 | GH | B.1.526 | Spike D614G, N M234I, N P168S, N P199L, NS3 P42L, NS3 Q57H, NS8 T11I, NSP2 T85I, NSP4 L438P, NSP5 P241S, NSP6 F108del, NSP6 G107del, NSP6 S106del, NSP12 P323L, NSP13 G54C |
| P21-2192 | 23 | Janssen | Y (COVID) | M | 41-50 | 25.3 | NY | New York | EPI_ISL_1790046 | NYULH1415 | GR | B.1.1.7 | Spike A570D, Spike D614G, Spike P681H, Spike T716I, N D3L, N G204R, N R203K, N S235F, NS8 K68stop, NS8 Q27stop, NS8 Y73C, NSP6 R138del, NSP6 V139del, NSP6 W140del |
| P21-2193 | 94 | Pfizer | N | F | 41-50 | 24.5 | NY | Nassau | EPI_ISL_1790047 | NYULH1416 | GR | P.1 | Spike D614G, Spike H655Y, Spike K417T, Spike T1027I, Spike V1176F, N G204R, N P80R, N R203K, NS3 S253P, NS8 E92K, NSP3 K977Q, NSP3 P822S, NSP6 F108del, NSP6 G107del, NSP6 K4R, NSP6 S106del, NSP12 P94L, NSP12 P323L, NSP13 E341D, NSP14 A119V |

| | | | | | | | | | | | | | |
|---|---|---|---|---|---|---|---|---|---|---|---|---|---|
| P21-2331 | 17 | Pfizer | Y (COVID) | F | 81-90 | 23.2 | NY | Queens | EPI_ISL_1922825 | NYULH1587 | GH | B.1.526 | Spike A701V, Spike D253G, Spike D614G, Spike E484K, Spike L5F, Spike M1229T, Spike T95I, N M234I, N P199L, NS3 P42L, NS3 Q57H, NS8 T11I, NSP2 T85I, NSP4 L438P, NSP6 F108del, NSP6 G107del, NSP6 S106del, NSP12 P323L, NSP13 H164Y, NSP13 Q88H |
| P21-2364 | 59 | Pfizer | N | M | 51-60 | 31.0 | NY | Kings | EPI_ISL_1922853 | NYULH1616 | GR | B.1.1.7 | Spike A570D, Spike D614G, Spike D1118H, Spike H69del, Spike P681H, Spike S982A, Spike T716I, Spike V70del, Spike Y144del, N D3L, N G204R, N R203K, N S235F, NS8 K68stop, NS8 Q27stop, NS8 R52I, NS8 Y73C, NSP3 A890D, NSP3 I1412T, NSP3 T183I, NSP3 T1127I, NSP6 F108del, NSP6 G107del, NSP6 S106del, NSP12 P227L, NSP12 P323L, NSP14 P451S |
| P21-2365 | 57 | Pfizer | N | F | 21-30 | 26.7 | NY | Kings | EPI_ISL_1922854 | NYULH1617 | GH | B.1.526.1 | Spike D80G, Spike D614G, Spike D950H, Spike F157S, Spike L452R, Spike T859N, Spike Y144del, N M234I, N T205I, NS3 P42L, NS3 P104L, NS3 Q57H, NS8 A51S, NS8 T11I, NSP2 T85I, NSP4 A446V, NSP4 K399E, NSP4 L438P, NSP5 F223I, NSP5 R222Q, NSP6 F108del, NSP6 G107del, NSP6 S106del, NSP6 V278I, NSP7 Q63R, NSP12 P323L |
| P21-2366 | 57 | Pfizer | N | M | 71-80 | 24.2 | NY | Kings | EPI_ISL_1922855 | NYULH1618 | GH | B.1.526.2 | Spike D253G, Spike D614G, Spike Q957R, Spike S477N, Spike T95I, N P13L, N S202R, NS3 P42L, NS3 Q57H, NS7a L116F, NS8 T11I, NSP2 T85I, NSP3 A1324T, NSP3 G1128S, NSP3 T169I, NSP4 L438P, NSP6 F108del, NSP6 G107del, NSP6 S106del, NSP9 D78N, NSP12 P227S, NSP12 P323L, NSP13 Q88H, NSP15 H337Y |
| P21-2368 | 52 | Pfizer | N | F | 21-30 | 22.0 | NY | New York | EPI_ISL_1922856 | NYULH1619 | GR | B.1.1.7 | Spike A570D, Spike D614G, Spike D1118H, Spike H69del, Spike K1191N, Spike P681H, Spike S982A, Spike T716I, Spike V70del, Spike Y144del, N D3L, N G204R, N R203K, N S235F, NS3 P240S, NS8 Q27stop, NS8 R52I, NS8 Y73C, NSP3 A890D, NSP6 F108del, NSP6 G107del, NSP6 S106del, NSP8 Q24R, NSP12 P323L, NSP13 K460R, NSP16 K160R, NSP16 T140I |
| P21-2370 | 48 | Pfizer | N | M | 71-80 | 31.2 | NY | New York | EPI_ISL_1922857 | NYULH1620 | GH | B.1.526 | Spike A701V, Spike D253G, Spike D614G, Spike E484K, Spike T95I, N M234I, N P199L, NS3 P42L, NS3 Q57H, NS8 T11I, NSP2 T85I, NSP4 L438P, NSP6 F108del, NSP6 G107del, NSP6 S106del, NSP12 P323L, NSP13 Q88H |
| P21-2371 | 54 | Moderna | N | F | 61-70 | 20.3 | NY | New York | EPI_ISL_1922858 | NYULH1621 | GH | B.1.429 | Spike D614G, Spike L452R, N T205I, NS3 Q57H, NSP2 T85I, NSP3 V1331F, NSP5 K90R, NSP9 I65V, NSP12 P323L, NSP13 D260Y, NSP14 S418C, NSP16 S33N |
| P21-2372 | 66 | Pfizer | N | M | 21-30 | 20.6 | NJ | Hudson | EPI_ISL_1922859 | NYULH1622 | GH | B.1.526 | Spike A701V, Spike D253G, Spike D614G, Spike T95I, N M234I, N P199L, NS3 P42L, NS3 P104L, NS3 Q57H, NS8 T11I, NSP2 A386S, NSP2 T85I, NSP3 S1588L, NSP4 L438P, NSP6 F108del, NSP6 G107del, NSP6 S106del, NSP12 P323L, NSP13 Q88H |
| P21-2373 | 78 | Pfizer | N | M | 31-40 | 33.1 | NY | New York | EPI_ISL_1922860 | NYULH1623 | G | B.1.1.7 | Spike A222S, Spike H69del, Spike V70del, Spike Y144del, M G89S, N D3L, N D401Y, NS3 L15F, NS8 S24L, NSP3 A890D, NSP4 F17L, NSP6 F108del, NSP6 G107del, NSP6 S106del, NSP13 K460R, NSP14 S171G, NSP16 A83T |
| P21-2375 | 105 | Pfizer | N | M | 51-60 | 30.3 | NY | Queens | EPI_ISL_1922861 | NYULH1624 | GH | B.1.526 | Spike A701V, Spike D253G, Spike D614G, Spike E484K, Spike T95I, N M234I, N P199L, NS3 P42L, NS3 Q57H, NS8 T11I, NSP2 T85I, NSP4 L438P, NSP6 F108del, NSP6 G107del, NSP6 S106del, NSP12 P323L, NSP13 Q88H |
| P21-2376 | 27 | Pfizer | N | F | 21-30 | 19.3 | NY | Nassau | EPI_ISL_1922862 | NYULH1625 | GH | B.1.526.1 | Spike D80G, Spike D614G, Spike F157S, Spike L452R, Spike T859N, Spike Y144del, N M234I, N T205I, NS3 P42L, NS3 P104L, NS3 Q57H, NS7a L116F, NS8 A51S, NS8 T11I, NSP2 T85I, NSP3 A861S, NSP4 A446V, NSP4 L438P, NSP6 F108del, NSP6 G107del, NSP6 S106del, NSP6 V278I, NSP12 P323L |
| P21-2378 | 20 | Pfizer | N | M | 61-70 | 18.1 | NY | Westchester | EPI_ISL_1922863 | NYULH1627 | GR | B.1.1 | Spike A222V, Spike D614G, Spike G769V, Spike S255F, N G204R, N Q418H, N R203K, N S187L, NSP2 A306V, NSP3 S1656A, NSP12 P323L, NSP13 G439R, NSP14 P412H |
| P21-2379 | 65 | Pfizer | N | F | 41-50 | 22.2 | NY | Rockland | EPI_ISL_1922864 | NYULH1628 | GR | B.1.1.7 | Spike A570D, Spike D614G, Spike D1118H, Spike H69del, Spike N148T, Spike P681H, Spike S982A, Spike T716I, Spike V70del, Spike Y144del, N D3L, N G204R, N R203K, N S235F, NS3 L15F, NS8 G8R, NS8 Q27stop, NS8 R52I, NS8 Y73C, NSP3 A890D, NSP3 S126L, NSP3 T183I, NSP4 F17L, NSP6 F108del, NSP6 G107del, NSP6 S106del, NSP12 P323L, NSP13 K460R |
| P21-2380 | 70 | Pfizer | N | M | 31-40 | 22.9 | NY | Bronx | EPI_ISL_1922865 | NYULH1629 | GH | B.1.526.1 | Spike D80G, Spike D614G, Spike D950H, Spike F157S, Spike L452R, Spike T859N, Spike Y144del, N M234I, N T205I, NS3 P42L, NS3 Q57H, NS8 A51S, NS8 F3L, NS8 T11I, NSP2 T85I, NSP4 A446V, NSP4 K399E, NSP4 L438P, NSP5 P96S, NSP6 F108del, NSP6 G107del, NSP6 S106del, NSP6 V278I, NSP12 P323L, NSP14 V398L |
| P21-2381 | 64 | Moderna | N | M | 71-80 | 22.1 | NY | Richmond | EPI_ISL_1922866 | NYULH1630 | GH | B.1.575 | Spike A1078S, Spike D614G, Spike P681H, Spike T716I, M I82T, N G96C, N K127R, N L224F, N T205I, NS3 Q57H, NSP2 S122F, NSP2 T85I, NSP3 A333S, NSP4 T492I, NSP6 T181I, NSP12 A97V, NSP12 P323L, NSP13 P47S |
| P21-2382 | 26 | Pfizer | N | M | 61-70 | 20.2 | NY | Nassau | EPI_ISL_1922867 | NYULH1631 | GR | B.1.1.7 | Spike A570D, Spike D614G, Spike D1118H, Spike H69del, Spike P681H, Spike S982A, Spike T716I, Spike V70del, Spike Y144del, N D3L, N G204R, N R203K, N S235F, NS8 C61F, NS8 K68stop, NS8 Q27stop, NS8 R52I, NS8 Y73C, NSP3 A890D, NSP6 F108del, NSP6 G107del, NSP6 S106del, NSP12 P227L, NSP12 P323L |
| P21-2383 | 17 | Pfizer | N | M | 41-50 | 32.7 | NY | New York | EPI_ISL_1922868 | NYULH1632 | GH | B.1.526 | Spike A701V, Spike D253G, Spike D614G, Spike E484K, Spike T95I, N M234I, N P199L, NS3 P42L, NS3 Q57H, NS8 T11I, NSP2 T85I, NSP4 L438P, NSP6 F108del, NSP6 G107del, NSP6 S106del, NSP12 M601I, NSP12 P323L, NSP13 Q88H |
| P21-2384 | 50 | Moderna | N | F | 71-80 | 24.5 | NY | Nassau | EPI_ISL_1922869 | NYULH1633 | GRY | B.1.1.7 | Spike A570D, Spike D614G, Spike D1118H, Spike H69del, Spike N501Y, Spike P681H, Spike S982A, Spike T716I, Spike V70del, Spike Y144del, N D3L, N G204R, N R203K, N S235F, NS3 P240S, NS8 K68stop, NS8 Q27stop, NS8 R52I, NS8 Y73C, NSP3 A890D, NSP3 E95D, NSP3 I1412T, NSP3 K963R, NSP3 M1441I, NSP3 T183I, NSP6 F108del, NSP6 G107del, NSP6 S106del, NSP12 P227L, NSP12 P323L |
| P21-2385 | 72 | Pfizer | N | M | 51-60 | 29.4 | NY | Nassau | EPI_ISL_1922870 | NYULH1634 | GRY | B.1.1.7 | Spike A570D, Spike D614G, Spike D1118H, Spike H69del, Spike N501Y, Spike P681H, Spike S982A, Spike T716I, Spike V70del, Spike Y144del, N D3L, N G204R, N R203K, N S235F, NS8 K68stop, NS8 Q27stop, NS8 R52I, NS8 Y73C, NSP3 A890D, NSP3 I310T, NSP3 I1412T, NSP3 M1441I, NSP3 P108L, NSP3 T183I, NSP6 F108del, NSP6 G107del, NSP6 S106del, NSP12 P227L, NSP12 P323L |
| P21-2388 | 31 | Pfizer | N | M | 31-40 | 18.5 | NY | Queens | EPI_ISL_1922872 | NYULH1636 | S | A.2.5.2 | Spike D215A, Spike D614G, Spike G142del, Spike ins215AGY, Spike L141del, Spike L452R, Spike V143del, N M234I, N P365S, N P383L, N S197L, NS3 S74F, NS7a A105S, NS8 L84S, NSP1 L4F, NSP3 K839E, NSP4 F308Y, NSP4 T492I, NSP6 H11Q, NSP12 M629T |
| P21-2535 | 108 | Pfizer | N | M | 51-60 | 14.4 | NY | Suffolk | EPI_ISL_2202736 | NYULH1755 | G | B.1.301 | Spike A688V, Spike D614G, Spike L18R, Spike N440K, Spike P681H, Spike R158S, Spike S735A, Spike T95I, Spike T1027I, N N213Y, N S194L, NS3 V256I, NS7b S5L, NS8 R52K, NSP2 A205V, NSP3 A1305V, NSP3 P1921S, NSP3 S1333I, NSP3 S1475G, NSP4 A380V, NSP4 T354A, NSP6 F108del, NSP6 G107del, NSP6 S106del, NSP12 G671S, NSP12 P323L, NSP13 P77L, NSP16 T110I |
| P21-2563 | 42 | Janssen | Y (not COVID) | F | 61-70 | 23.1 | NY | Nassau | EPI_ISL_2161757 | NYULH1782 | GH | B.1.429 | Spike D614G, Spike L452R, Spike S13I, Spike W152C, N M234I, N T205I, NS3 Q57H, NS8 V100L, NSP2 A510V, NSP2 E574G, NSP2 F300L, NSP2 R46K, NSP2 T85I, NSP2 Y441C, NSP9 I65V, NSP12 P323L, NSP13 D260Y |
| P21-2567 | 77 | Pfizer | N | F | 71-80 | 19.6 | NY | New York | EPI_ISL_2161761 | NYULH1786 | GRY | B.1.1.7 | Spike A570D, Spike D614G, Spike D1118H, Spike H69del, Spike N501Y, Spike P681H, Spike S982A, Spike T716I, Spike V70del, Spike V327I, Spike Y144del, N D3L, N G204R, N P67S, N R203K, N S235F, NS3 A110S, NS8 K68stop, NS8 Q27stop, NS8 R52I, NS8 Y73C, NSP3 A890D, NSP3 I1412T, NSP3 T183I, NSP6 F108del, NSP6 G107del, NSP6 S106del, NSP12 P227L, NSP12 P323L, NSP14 P451S |

| | | | | | | | | | | | | | |
|---|---|---|---|---|---|---|---|---|---|---|---|---|---|
| P21-2573 | 100 | Pfizer | N | F | 31-40 | 19.3 | NY | Queens | EPI_ISL_2161767 | NYULH1792 | GRY | B.1.1.7 | Spike A570D, Spike D614G, Spike D1118H, Spike H69del, Spike N501Y, Spike P681H, Spike S982A, Spike T716I, Spike V70del, Spike V327I, Spike Y144del, N D3L, N G204R, N R203K, N S235F, NS8 K68stop, NS8 Q27stop, NS8 R52I, NS8 Y73C, NSP3 A890D, NSP3 I1412T, NSP3 T183I, NSP6 F108del, NSP6 G107del, NSP6 S106del, NSP12 P227L, NSP12 P323L, NSP14 P451S |
| P21-2577 | 92 | Pfizer | N | F | 31-40 | 25.3 | NY | New York | EPI_ISL_2202787 | NYULH1796 | GRY | B.1.1.7 | Spike A570D, Spike D614G, Spike D1118H, Spike H69del, Spike N501Y, Spike P681H, Spike S982A, Spike T716I, Spike V70del, Spike Y144del, N D3L, N E367D, N G204R, N R203K, N S235F, NS3 P240S, NS7a C113L, NS7a F114H, NS7a I110N, NS7a L116Q, NS7a R118K, NS8 K68stop, NS8 Q27stop, NS8 R52I, NS8 Y73C, NSP3 A890D, NSP3 E95D, NSP3 I1412T, NSP3 M1441I, NSP3 T183I, NSP6 F108del, NSP6 G107del, NSP6 L260F, NSP6 S106del, NSP12 P227L, NSP12 P323L |
| P21-2580 | 23 | Pfizer | N | M | 61-70 | 27.5 | NY | Suffolk | EPI_ISL_2161773 | NYULH1799 | GH | B.1.2 | Spike A570V, Spike D614G, Spike G142V, N P67S, N P199L, NS3 G172V, NS3 Q57H, NS3 T24N, NS8 S24L, NSP1 G82del, NSP1 H83del, NSP1 M85del, NSP1 V84del, NSP1 V86del, NSP2 G265del, NSP2 L266del, NSP2 T85I, NSP3 T3A, NSP5 L89F, NSP12 P323L, NSP12 V637I, NSP14 N129D, NSP16 R216C |
| P21-2592 | 99 | Pfizer | N | F | 21-30 | 25.4 | NY | Kings | EPI_ISL_2161781 | NYULH1811 | GH | B.1.526 | Spike A701V, Spike D253G, Spike D614G, Spike T95I, N M234I, N P199L, NS3 P42L, NS3 P262S, NS3 Q57H, NS8 T11I, NSP2 T85I, NSP3 P1469S, NSP3 S1396T, NSP4 L438P, NSP6 F108del, NSP6 G107del, NSP6 S106del, NSP12 P323L, NSP13 Q88H |
| P21-2594 | 102 | Pfizer | N | F | 31-40 | 18.2 | NY | New York | EPI_ISL_2161782 | NYULH1812 | G | B.1 | Spike D614G, Spike H655Y, N T205I, NS3 S253P, NS3 V256F, NS8 E92K, NSP6 F108del, NSP6 G107del, NSP6 S106del, NSP13 E341D |
| P21-2738 | 49 | Moderna | N | M | 71-80 | 24.2 | NY | Nassau | EPI_ISL_2400287 | NYULH1814 | GRY | B.1.1.7 | Spike A570D, Spike D614G, Spike D1118H, Spike H69del, Spike K1191N, Spike N501Y, Spike P681H, Spike S982A, Spike T716I, Spike V70del, Spike Y144del, N D3L, N G204R, N R203K, N S235F, NS8 Q27stop, NS8 R52I, NS8 Y73C, NSP3 A225V, NSP3 A890D, NSP3 I1412T, NSP3 T183I, NSP6 F108del, NSP6 G107del, NSP6 S106del, NSP8 Q24R, NSP12 P323L, NSP13 K460R, NSP14 D324E, NSP16 K160R |
| P21-2764 | 27 | Pfizer | Y (COVID) | F | 51-60 | 30.8 | NY | New York | EPI_ISL_2400336 | NYULH1815 | GRY | B.1.1.7 | Spike A570D, Spike D614G, Spike D1118H, Spike H69del, Spike N501Y, Spike P681H, Spike S982A, Spike T716I, Spike V70del, Spike Y144del, E S68F, N D3L, N G204R, N R203K, N S235F, NS3 D210Y, NS8 K68stop, NS8 Q27stop, NS8 R52I, NS8 Y73C, NSP3 A890D, NSP3 I1412T, NSP3 M1441I, NSP3 T183I, NSP6 F108del, NSP6 G107del, NSP6 S106del, NSP12 P227L, NSP12 P323L, NSP13 A509V |
| P21-2765 | 28 | Moderna | N | M | 81-90 | 17.2 | NY | Nassau | EPI_ISL_2400382 | NYULH1816 | GH | B.1.526 | Spike A701V, Spike D253G, Spike D614G, Spike L5F, Spike T95I, N M234I, N P199L, NS3 P42L, NS3 Q57H, NS8 T11I, NSP2 T85I, NSP4 L438P, NSP5 T21I, NSP6 F108del, NSP6 G107del, NSP6 L37F, NSP6 S106del, NSP12 P323L, NSP13 Q88H |
| P21-3585 | 98 | Pfizer | Y (not COVID) | M | 61-70 | 36.0 | NY | Queens | EPI_ISL_3066854 | NYULH2344 | G | AV.1 | Spike D614G, Spike D1118H, Spike G142D, Spike K1191N, Spike P681R, Spike S982A, Spike T716I, N R203M, NS8 Q27stop, NS8 R52I, NSP3 P654S, NSP4 R401H, NSP6 F108del, NSP6 G107del, NSP6 S106del, NSP12 G671S, NSP12 P323L |
| P22-1140 | 23 | Pfizer | Y (COVID) | M | 61-70 | 14.7 | NY | New York | EPI_ISL_1322825 | NYULH877 | GH | B.1.526.2 | Spike D253G, Spike D614G, Spike Q957R, Spike S477N, Spike T95I, N A414V, N P13L, N S202R, NS3 P42L, NS3 Q57H, NS7a L116F, NS8 T11I, NSP2 T85I, NSP3 G1128S, NSP3 T1022I, NSP4 L438P, NSP6 F108del, NSP6 G107del, NSP6 S106del, NSP12 P323L, NSP13 Q88H, NSP13 V521I |

Supplementary Table 4: Spike mutation statistics at 23 sites with enriched mutation rates in vaccine breakthrough infections in New York.

| Spike_pos | 5 | 69 | 70 | 95 | 142 | 144 | 157 | 215 | 222 | 327 | 417 | 477 | 484 | 501 | 570 | 655 | 681 | 716 | 950 | 982 | 1027 | 1118 | 1191 |
|---|---|---|---|---|---|---|---|---|---|---|---|---|---|---|---|---|---|---|---|---|---|---|---|
| hCoV-19/Wuhan-Hu-1 | L | H | V | T | G | Y | F | D | A | V | K | S | E | N | A | H | P | T | D | S | T | D | K |
| spike mutation | L5X | H69X | V70X | T95X | G142X | Y144X | F157X | D215X | A222X | V327X | K417X | S477X | E484X | N501X | A570X | H655X | P681X | T716X | D950X | S982X | T1027X | D1118X | K1191X |
| important sites | | del | | | | del | | | | | RBD (aa 331 - 524) | | | | RBD up/down | | S1/S2 cleavage | | | | | | |
| Unvacc-ctr WT (%) | 62.82 | 69.46 | 70.71 | 65.99 | 99.11 | 59.69 | 89.6 | 99.42 | 99.23 | 99.76 | 98.5 | 89.24 | 76.96 | 69.04 | 71 | 98.76 | 61.82 | 68.16 | 90 | 70.91 | 98.85 | 70.76 | 98.05 |
| Unvacc-ctr mut (%) | 37.18 | 30.54 | 29.29 | 34.01 | 0.89 | 40.31 | 10.4 | 0.58 | 0.77 | 0.24 | 1.5 | 10.76 | 23.04 | 30.96 | 29 | 1.24 | 38.18 | 31.84 | 10 | 29.09 | 1.15 | 29.24 | 1.95 |
| Vacc WT (%) | 60 | 66.67 | 66.67 | 63.89 | 95.45 | 53.03 | 89.23 | 97.18 | 97.14 | 95.83 | 96.83 | 88.89 | 68.89 | 66.67 | 63.16 | 97.33 | 54.67 | 61.84 | 89.66 | 65.33 | 96.05 | 65.28 | 95.77 |
| Vacc mut (%) | 40 | 33.33 | 33.33 | 36.11 | 4.55 | 46.97 | 10.77 | 2.82 | 2.86 | 4.17 | 3.17 | 11.11 | 31.11 | 33.33 | 36.84 | 2.67 | 45.33 | 38.16 | 10.34 | 34.67 | 3.95 | 34.72 | 4.23 |
| Δ (Vacc mut % minus Unvacc mut %) | 2.82 | 2.79 | 4.04 | 2.1 | 3.66 | 6.66 | 0.37 | 2.24 | 2.09 | 3.93 | 1.67 | 0.35 | 8.07 | 2.37 | 7.84 | 1.43 | 7.15 | 6.32 | 0.34 | 5.58 | 2.8 | 5.48 | 2.28 |
| p (Fisher) | 0.74 | 0.61 | 0.51 | 0.70 | 0.03 | 0.30 | 0.84 | 0.09 | 0.13 | 0.02 | 0.27 | 0.82 | 0.21 | 0.74 | 0.15 | 0.27 | 0.22 | 0.26 | 0.82 | 0.30 | 0.08 | 0.35 | 0.18 |
| q (multiplicity correction, BH) | 0.84 | 0.82 | 0.74 | 0.84 | 0.37 | 0.50 | 0.84 | 0.50 | 0.50 | 0.37 | 0.50 | 0.84 | 0.50 | 0.84 | 0.50 | 0.50 | 0.50 | 0.50 | 0.84 | 0.50 | 0.50 | 0.54 | 0.50 |

**Supplementary Methods**

**cDNA synthesis, library preparation and sequencing**

Total RNA (11 µl) was converted to first strand cDNA by random priming using the Superscript IV first-strand synthesis system (Invitrogen, ref# 180901050). Random priming was performed as follows: 65˚C for 5 mins, cooled on ice followed by addition of mix to run cDNA first strand synthesis 23˚C 10mins, 50˚C for 1 hour, 80˚C for 10 mins, and 4˚C hold. Libraries were prepared using Swift Normalase Amplicon Panel (SNAP) SARS-CoV-2 and Sar-Cov-2 additional Genome Coverage (Cat# SN-5X296 core kit, 96rxn), using 10 µl of first strand cDNA, following the manufacturer's instruction: https://swiftbiosci.com/wp-content/uploads/2021/06/PRT-028-Swift-Normalase-Amplicon-Panels-SNAP-SARS-CoV-2-Panels-Rev-9.pdf). First step incorporated tiled primer pairs which target and enrich for the entire 29.9kb covid-19 viral genome (NCBI Reference Sequence NC_045512.2) during multiplex PCR. Multiplex PCR was run as follows: 98˚C 30 secs, 4 repeating cycles of 98˚C/ 10secs, 61˚C/5mins, 65˚C/1min. 24 repeating cycles of 98˚C/10secs, 64˚C/1min followed by 1 cycle of 65˚C for 1 min and held at 4 ˚C. The multiplexed samples were cleaned with 1x volume room temperature Ampure XP beads (Beckman Coulter, #A63882) for 5 mins, washed with 80% ethanol twice, and beads were resuspended in 17.4 µl of TE buffer (part of swift kit). Indexing reagent mix was added to the resuspended beads. One unique SNAP UD dual indexing primer pair was added to each sample. Indexing PCR was run at 37˚C/20mins, 98˚C/30secs, 9 repeating cycles of 98˚C/10secs, 60˚C/30secs, 66˚C/1min. After indexing PCR was completed, samples were cleaned with 32.5ul of PEG NaCl solution (provided in swift kit, 5min bind time, 80% ethanol wash x2) and eluted in 20ul of post PCR TE buffer. Final libraries were run on Agilent Tapestation 2200 with high sensitivity DNA Screentape to verify amplicon size of about 450 bp. Samples were pooled according to the enzymatic normalase step. The loading concentration for the normalized pool was verified on QPCRusing the Kapa library quantification complete kit (Kapa-Roche, #KK4824) run on the Bio-Rad CFX384 Real-time system. Normalized pools were run on the Illumina NovaSeq 6000 system with the SP 300 cycle flow cell. Run metrics were paired end 150 cycles with dual indexing reads. Typically, 2 pools representing 2 full 96 well plates (192 samples) were sequenced on each SP300 NovaSeq flow cell.

**Sequenced read processing**

Binary base call files outputted by the Illumina NovaSeq 6000 sequencing instrument (RTA Version 3.4.4) were converted to FASTQ format and demultiplexed according to the unique dual-index adapter sequences of each sample using the Illumina bcl2fastq2 Conversion Software v2.20. Adapters and low quality bases were trimmed using Trimmomatic v0.36 (Bolger et al. 2014) with the settings 'ILLUMINACLIP:trimmomatic.fa:2:30:10:1:true TRAILING:5 SLIDINGWINDOW:4:15 MINLEN:35'. The BWA-MEM algorithm of the alignment program, BWA v0.7.17 (Li and Durbin 2009), was utilized for mapping reads of each sample to the SARS-CoV-2 reference genome (NC_045512.2, wuhCor1) with the '-M' flag for marking shorter split hits as secondary and the parameter '-U' set to a 17 alignment score penalty for an unpaired read pair. Name-sorted mapped reads in SAM alignment format were soft-clipped to remove primer sequences specific to the short overlapping (tiled) amplicon design of the Swift Normalase

Amplicon SARS-CoV-2 Panel using Primerclip v0.3.8 (Swift Biosciences: https://github.com/swiftbiosciences/primerclip), and the Picard v2.18.20 “AddOrReplaceReadGroups” tool (Broad Institute: http://broadinstitute.github.io/picard/) was used in converting the primer-trimmed SAM files to coordinate-sorted and indexed BAM format with read-groups added for downstream analysis. The variant caller, BCFtools v1.9 (Li et al. 2009), was utilized to detect genetic mutations of the collected viral samples from BAM input:

bcftools mpileup -r NC_045512.2 --count-orphans --no-BAQ --max-depth 50000 --max-idepth 500000 --annotate FORMAT/AD,FORMAT/ADF,FORMAT/ADR,FORMAT/DP,FORMAT/SP,INFO/AD,INFO/ADF,INFO/ADR --output-type v |
bcftools call --ploidy 1 --keep-alts --multiallelic-caller --output-type v |
bcftools norm --multiallelics -any --output-type v |
bcftools filter -i "(DP4[0]+DP4[1])<(DP4[2]+DP4[3]) && ((DP4[2]+DP4[3])>0) & FORMAT/AD[0:1]>20" --output-type v |
bcftools filter -e "IMF<0.5" --output-type v

To generate viral sequences, the output in VCF format was applied to the reference sequence using `bcftools consensus` while the BEDTools v2.30.0 utilities (Quinlan and Hall 2010), “genomecov” (with the ‘-bga’ option to report depth at each base including zero coverage sites), “merge” (for creating a BED file of low depth sites with <1000x coverage to be masked), and “maskfasta” in conjunction with the multiple sequence alignment program, MAFFT v4.471 (Katoh et al. 2002) in ‘--auto’ mode, were used in the production of final consensus sequences with all bases below 1000x coverage masked and both non-targeted ends of the sequences trimmed. Sequences with fewer than 23,000 bp with at least 4000x coverage were discarded from further analysis for having not yielded a near-complete SARS-CoV-2 genome, resulting in 1,254 final sequences suitable for downstream analysis (Supplemental Table). Each assembled SARS-CoV-2 genome sequence was assigned the most likely phylogenetic lineage according to Phylogenetic Assignment of Named Global Outbreak Lineages (PANGO) nomenclature (Rambaut et al. 2020) using the PANGO lineage tool (version: 2021-06-05).
